# Mental Health Symptom Reduction Using Digital Therapeutics Care Informed by Genomic SNPs and Gut Microbiome Signatures

**DOI:** 10.1101/2022.04.27.22273901

**Authors:** Inti Pedroso, Shreyas V. Kumbhare, Bharat Joshi, Santosh K. Saravanan, Dattatray S. Mongod, Simitha Singh-Rambiritch, Tejaswini Uday, Karthik M. Muthukumar, Carmel Irudayanathan, Chandana Reddy-Sinha, Parambir S. Dulai, Ranjan Sinha, Daniel E. Almonacid

**Affiliations:** Digbi Health, Mountain View, California, USA; National Centre for Cell Science, Pune, Maharashtra, India; Division of Gastroenterology, Northwestern University, Chicago, Illinois, USA

**Keywords:** Anxiety, Depression, Insomnia, Gut-brain-axis, Multi-omic models, Non-pharmacological treatment

## Abstract

**Background:** Mental health diseases are a major component of morbidity and health care costs. Obesity and gut disorders are comorbid with mental health, with the gut microbiome hypothesized to play a key mechanistic role in linking them. Pharmacological and behavioral interventions are currently used to treat mental health disorders, but they have limited efficacy. Dietary and weight-loss interventions have been shown to provide different benefits. Still, there exists conflicting evidence regarding their effects which may be due to an individual’s genetic or microbiome factors modulating the improvement of symptoms.

**Objective:** We aimed to identify genetic and gut microbiome factors that explain the improvement in mental health after a dietary and lifestyle intervention for weight loss.

**Methods:** We recruited 369 individuals participating in the Digbi Health personalized digital care program, for which we evaluated the association between 23 genetic scores, the abundance of 178 gut microbiome genera, and 42 gut-brain modules (pathways related to neuroactive metabolites produced by gut microbes) with the presence/absence of anxiety/depression or sleep problems at baseline and improvement on anxiety, depression, and insomnia after losing at least 2% body weight.

**Results:** The mean BMI and age of the study cohort were 34.6 and 48.7, respectively, and there was an overrepresentation of individuals with functional gastrointestinal disorders (84%). On average, the individuals lost 5.4% of body weight at the time of follow-up (mean of 88 days), and more than 95% reported improvement in at least one outcome. We found significant correlations between genetic scores with anxiety and depression at baseline, gut microbial functions with sleep problems at baseline, and genetic scores and gut microbial taxa and functions with anxiety, depression, and insomnia improvement. Among the gut microbial functions identified, the abundance of butyrate synthesis genes was associated with less than average improvement in depression symptoms, the abundance of kynurenine synthesis genes was associated with less than average improvement in anxiety symptoms, and the abundance of genes able to synthesize and degrade neuroactive hormones like nitric oxide was associated with greater than average improvement in depression and insomnia symptoms. Among the genetic scores identified, anxiety or depression at baseline were associated with genetic scores for alcohol use disorder and major depressive disorder, and greater than average improvement in anxiety and depression symptoms was associated with an obstructive sleep apnea genetic score. Furthermore, a type 1 diabetes genetic score was associated with a greater than average improvement of insomnia symptoms, whereas a type 2 diabetes genetic score was associated with a less than average improvement of insomnia symptoms. We compared the relative ability of demographic, genetic, and microbiome factors to explain baseline and improvement in mental health and found that genetic and microbiome factors provide value above demographic variables alone. Medication and recreational drug use do not confound microbiome associations with mental health.

**Conclusions:** The digital therapeutics care program significantly decreased body weight and concomitantly decreased self-reported mental health symptom intensity. Our results provide evidence that genetic and gut microbiome factors help explain interindividual differences in mental health improvement after dietary and lifestyle interventions for weight loss. Thus, individual genetic and gut microbiome factors provide a basis for designing and further personalizing dietary interventions to improve mental health.

## Introduction

Poor mental health is a significant determinant of health-related quality of life with important implications at individual and population levels. Pharmacological and behavioral interventions prevent and treat individuals at risk of or suffering from mental health disorders, but their efficacy is limited, and many experiencing improvements will relapse [1]. The COVID19 pandemic resurfaced and worsened the mental health crisis and brought awareness to society of its relationship with obesity and other chronic health conditions [2]. There is a great need to develop cost-effective interventions that provide significant short and long-term therapeutic benefits to individuals suffering from mental health, especially those with multiple comorbid conditions. Digital therapeutics have gained increased attention as a strategy to provide care to large numbers of individuals, and emerging evidence suggests its effectiveness for several chronic diseases [3, 4].

The epidemiological literature provides compelling evidence indicating that genetic and non-genetic factors contribute to the etiology of mental health disorders [5–7]. Additionally, substantial evidence exists linking mental health with digestive and gut disorders. For instance, two meta-analyses found a higher rate of anxiety and depression among IBS patients [8, 9], and a recent genetic study identified genetic factors linking IBS with mental health disorders [10]. The increasing prevalence of mental health has been linked to the increased rate of obesity and lifestyle risk factors. A meta-analysis found a bidirectional relationship between the two, with depressed individuals having a 37% increased risk of being obese. Those who were obese had an 18% increased risk of being depressed [11]. Furthermore, human genetic studies have shown a causal link between higher BMI and depression [12, 13] and higher BMI and anxiety, among other psychological and psychiatric disorders [14].

Recently, extensive epidemiological studies coupled with gut microbiome sequencing are reinforcing the importance of the gut-brain axis and identifying the gut microbiome factors underlying it. A gut microbiome and behavioral study found associations between the gut microbiome’s taxa and functions with depression status and quality of life [15]. There has been growing evidence, including clinical trials focused on understanding probiotics’ effects, suggesting bidirectional gut-brain-axis communication in mental health [16].

From an intervention point of view, the current evidence shows that inter-individual variation in the gut microbiome composition is mainly due to non-genetic factors, i.e., diet, exercise, and medication [17] and, therefore, the gut microbiome primarily contributes to the non-genetic etiology of mental health disorders. However, exceptions to this pattern exist [18, 19]. Thus, modulation of the gut microbiome by pre- and pro-biotics and other dietary or lifestyle interventions is an essential avenue for preventing and treating mental health and other conditions (for instance, see [20]). There is mixed evidence regarding the effect of weight loss on mental health, with reports providing contradictory evidence [21–25]. It is plausible that interindividual differences, mediated by genetic or non-genetic factors such as the gut microbiome, can explain why some individuals improve their mental health after weight loss and others do not.

Digbi Health has implemented and commercialized a personalized digital care intervention that uses its multi-omics platform to provide dietary and lifestyle recommendations personalized using genetic and microbiome information. The intervention has been shown to provide body weight loss in over 70% of individuals [26], reduction of fasting blood glucose level by 17.55% and an average reduction of HbA1c level by 6.27% [27], and a significant reduction in the symptomatology of Functional Gastro-Intestinal Disorders (FGIDs) [28]. This study addresses if gut microbiome taxa or functions and genetic markers explain body weight loss’s effect on mental health. We focused our analysis on 369 overweight and obese participants on Digbi Health personalized digital care dietary and lifestyle intervention who reported baseline and follow-up mental health outcomes on depression, anxiety, and insomnia. Our results provide evidence for inherited genetics and gut microbiome factors predisposing individuals to improving mental health after weight loss. They offer an opportunity to personalize and develop tailored dietary recommendations to tackle obesity and mental health disorders.

## Methods

### Ethics approval

E&I Review Services, an independent institutional review board, reviewed and approved IRB Study #18053 on 05/22/2018. Additionally, IRB Study #21141 was determined to be exempt from IRB review by E&I Review Services on 08/06/2021. Research material derived from human participants included self-collected buccal and fecal swabs. Informed consent was obtained electronically from study participants.

### Study rationale

This study aimed to identify genetic and gut microbiome factors associated with mental health improvement after a dietary and lifestyle intervention. We included the study subjects that reported mental health conditions, particularly anxiety or depression when initiating the intervention. Study subjects were asked to rate their improvement in mental health after they had achieved at least 2% body weight loss through the digital therapeutic program. This low body weight loss threshold allowed us to have a broad range of body weight loss in this cohort. Additionally, our previously published research indicated that two thirds of high BMI individuals lost at least 2% body weight 120 days after initiating the intervention. As genetic information, we considered genetic scores for traits that are known comorbidities of anxiety, depression, and obesity (see details below). We added to the outcomes of the study sleep problems at baseline and insomnia at follow-up. Both are integral parts of anxiety and depression and are known to be associated with digestive and gut issues, like functional gastrointestinal disorders. As for the gut microbiome, we considered taxonomic annotation summarized at the level of genera and predicted functional pathways previously linked with mental health (see details below).

### Participant enrollment, intervention, and phenotype data collection

Study participants were recruited from February 2020 to October 2021 among those who achieved 2% or more body weight loss from the date when enrolled in the Digbi Health personalized digital care program. The study participants were provided with an online questionnaire that included questions regarding anxiety or depression at baseline as well as sleep problems at baseline. Those who indicated a positive answer to having anxiety or depression were asked to rate the intensity of their anxiety and depression on a scale from 0 (minimum) to 5 (maximum). Those that indicated a positive answer about having sleep problems at baseline were asked to rate the intensity of their insomnia on a scale from 0 (minimum) to 5 (maximum). Additionally, as part of the enrollment process for Digbi’s program study, participants provided information on the presence or absence of symptoms associated with FGIDs, prescribed and over-the-counter medications or supplements, alcohol intake, recreational drug usage, and demographic information, including age, gender, and height. Body mass index at baseline and follow-up was calculated using the closest weight measurement to the enrollment date and date on which the survey was answered (within ± 14 days). Individuals were classified as having an FGID if they reported one or more of the following seven conditions: constipation, diarrhea, gassiness, bloating, heartburn/acid reflux, abdominal pain (cramping/belly pain) and IBS [29, 30].

To identify those participants consuming either antidepressants, anti-anxiolytics, antibiotics, or antimycotics, we mapped the subjects’ reported medications names to the chembl database using the database API [31]. We further obtained the ATC codes (https://www.whocc.no/atc_ddd_index/) from which we identified medications prescribed for depression (N06AA, N06AB, and N06AX), anxiety (N05B) and antibiotics and antimycotics (J01B, J01C, J01D, J01E, J01F, J01G, J01M, J01R, J01X, J02, and J04).

Digbi Health’s intervention has been described elsewhere [28]. In a nutshell, personalization of plans is achieved by analyzing participants’ genetics, gut bacteria, lifestyle, and demographics. Based on these data, the program encourages participants to make incremental lifestyle changes focused on reducing sugar consumption and timing meals to optimize insulin sensitivity, reduce systemic inflammation by identifying possibly inflammatory and anti-inflammatory nutrients, and increase fiber diversity to improve gut health. Behavioral changes are implemented with the help of virtual health coaching and the app, ensuring they are habit-forming.

### Sample collection and processing: Genome SNP array and gut microbiome sequencing

Subjects self-collected saliva samples using buccal swabs (Mawi Technologies iSwab DNA collection kit, Model no. ISWAB-DNA-1200) and fecal samples using fecal swabs (Mawi Technologies iSWAB Microbiome collection kit, Model no. ISWAB-MBF-1200). Sample collection was completed by following standardized directions provided to all subjects in an instruction manual. Saliva DNA extraction, purification, and genotyping using Affymetrix’s Direct to Consumer Array version 2.0 (“DTC”) on the Affymetrix GeneTitan platform was performed by Akesogen Laboratories in Atlanta, GA. Genotype calling was performed using Axiom Analysis Suite Software version 5.1.1. Genotypes were set to ‘missing’ if the sample did not meet the confidence threshold for making the call or if the probeset did not fall into a recommended category. Specific details about the probeset metrics can be found in the Axiom Analysis Suite User Guide or the Genotyping Data Analysis Guide (ThermoFisher Scientific 2022). Sample processing of baseline (pre-intervention) fecal samples was followed by the 16S rRNA gene amplicon sequencing performed at Akesogen Laboratories in Atlanta, GA. DNA extraction was performed using Qiagen MagAttract Power Microbiome DNA Kit on an automated liquid handling DNA extraction instrument.

### Microbiome data analyses

The V3-V4 region of the 16S rRNA gene was amplified and sequenced on the Illumina MiSeq platform using 2 x 300 bp paired-end sequencing [32]. Sequence reads were demultiplexed, and ASVs generated using DADA2 in QIIME2 version 2020.8 [33]. Primers and low-quality bases (Q <30) were trimmed off the reads from forward and reverse reads. Taxonomic assignment was performed using the vsearch consensus method against the 99% non-redundant Silva 138 reference database [34]. We excluded hits to Mitochondria, Chloroplast, Eukaryota, and unassigned taxa at the phylum level [35]. Six samples for participants who reported antibiotic consumption were excluded from the downstream analysis. Any ASV not seen more than once in at least 10% of the samples were filtered out to remove ASVs with a small mean and trivially large coefficient of variation [35]. The abundance matrix was rarefied at even depth (n=36,000 reads per sample with 500 iterations) using the ‘q2-repeat-rarefy’ plugin from QIIME2 [36]. The abundance matrix was then agglomerated at the genus level, resulting in 178 taxa across 344 samples. We further filtered any taxa with read counts less than 10. Furthermore, the raw abundance data were subjected to centered-log ratios (CLR) transformation [37].

The microbial functional prediction was performed using the q2-picrust2 plugin (version 2021.2) in QIIME2 [38]. The abundance matrix comprising KEGG Orthologs was then used to obtain the abundance of specific pathways related to neuroactive metabolites produced by gut microbes, described as ‘Gut-Brain Modules’ (GBMs) in an earlier study [15]. Predicted gene abundance encoding these metabolites was derived using the Omixer-RPM package (version 0.3.2) [39].

### Genetic data analyses

Probe level genotype calls were unified at the variant level and formatted in VCF format, and multiallelic sites with discordant genotype calls among different probes were set as missing. VCF files were left normalized with bcftools version 1.14 (using htslib version 1.14). Beagle version 5.3 [40, 41] was used for phasing and imputation using the 1KG project as reference [42]. Processing of the 1KG data for imputation included only SNPs and Indels, removing sites with allele counts <2 and left normalization with bcftools, and converting filtered and phased VCF files to bref3 format using the bref3.28Jun21.220.jar script provided by the Beagle software suite. We included 13,478,023 variants on downstream analyses deriving from sites imputed with r^2^>=0.8 or chip-genotyped. All analyses were orchestrated by a SnakeMake pipeline [43].

We merged our un-imputed genotype data with the 1KG data, including only sites with MAF > 1% and genotype missingness < 10%. We performed PCA analysis and calculated the first 20 PCs using Plink2 [44] on the combined dataset. We focused on the main five super populations defined by the 1KG project to estimate the sample’s ancestry composition. We trained a random forest classifier as implemented on scikit-learn [45] with max depth = 8 and number of trees = 100 using 75% and 25% of the dataset for training and test, respectively. We obtained a Matthew’s correlation coefficient of 0.98 showing good performance as has been reported elsewhere for this strategy [46].

Twenty-three traits were selected based on being digestive system traits comorbid with mental health disorders, anxiety and sleep in particular, or mental health disorders that are known comorbidities of digestive system conditions focused on IBS, IBD, and obesity [47, 48]. We reviewed each trait’s genetic and genome-wide association studies and extracted summary statistics, including chromosome, position, effect allele, effect size, and ancestry of discovery and replication populations. Table S1 provides a detailed description of each trait and associated references as data sources. Genetic scores were calculated by multiplying the beta value or logarithm of the odds ratio by the number of risk alleles each individual had and the mean overall genetic variants included on the panel. All genetic scores were coded to be interpreted such that a larger genetic score is associated with increasing inherited genetic predisposition to the condition.

### Statistical analysis

To identify genetic and microbiome predictors associated with each outcome, we performed univariate analyses utilizing a multiple regression with demographic variables. We used logistic regression for sleep problems at baseline and anxiety or depression at baseline. We used Poisson regression to improve intensity scores, with the outcome being the intensity at T1 and offset being the intensity at T0. In all models, we included as demographic variables FGID: binary, the self-reported status of the previous diagnosis of a functional gastrointestinal disorder; gender: binary, male or female; BMI at T0: continuous variable; Age: continuous variable; weight loss: categorical variable, categorized as those with no change, lost 0 to 5%, 5-10% or more than 10% of their body weight at T1 in relation to T0; and five principal components (continuous variable) calculated using the genetic ancestry analyses described previously. Linear regression was fitted using the statsmodels python package [49] v0.13.2 using the Binomial and Poisson families with the identity and links for logistic and Poisson regression, respectively, and with the HC3 covariance matrix as suggested earlier [50]. These regression models identified microbiome and genetic factors measured at baseline that are associated with increased prevalence of mental health at baseline and different levels of improvement in mental health at follow-up. On the logistic models (linear models with Binomial family link), a regression coefficient greater than zero is interpreted as an increasing prevalence of self-reported illness with an increasing abundance of microbiome factors or a higher value of the genetic scores. On the Poisson regression models, a regression coefficient greater than zero is interpreted as a higher abundance of microbiome factors or a higher value of genetic scores being associated with less than average improvement in the outcome.

We applied correction for multiple hypothesis tests using the False Discovery Rate (FDR) [51] as implemented on the function “multiple tests” of the statsmodels python package and selected statistically significant results with an FDR ≤ 0.15.

We performed model comparisons between four different models by building regression models for demographic predictors only (D), demographic and microbiome predictors (D+M), demographic and genetic predictors (D+M), and all three sets of predictors (D+M+G). For microbiome and genetic predictors, we included in the models’ variables identified on univariate analyses with an FDR ≤ 0.15. We performed a singular value decomposition (SVD) analysis with the microbiome variables, bacterial genera, and functional pathways to avoid collinearity in the regression models. We used the singular vectors as predictors on the regression models. We selected the same number of singular vectors as the number of microbiome variables used for the SVD. We report adjusted pseudo r-squared values, which allow comparison between models with a different number of predictors; in particular, we used Pratt’s method [52], which is recommended for cases where the ratio between the number of samples and predictors is less than 100 [53]. Model comparisons were performed using Cox-Snell pseudo r-squared adjusted using Pratt’s methods to correct for the different number of predictors in the different models. To estimate the mean, median, standard deviation, and percentiles of the pseudo r-squared values, we performed bootstrap with 1001 bootstrap replicates. Bootstrap values were bias-corrected by subtracting the absolute value of the difference between the mean pseudo r-squared of the 1001 bootstrap replicates and the value obtained from the original non-bootstrapped data. This ensures that the mean of the bootstrap distributions is the same as the pseudo r-squared obtained from the original non-bootstrapped data. The bootstrap analysis is not meant for hypothesis testing but to provide uncertainty estimated around the pseudo r-squared reported.

To test the effect of potential confounders, particularly mental health medications and demographics, on the association of gut microbiome with mental health outcomes, we performed a multivariate analysis using PERMANOVA with 999 bootstrap iterations based on Bray-Curtis dissimilarity with the vegan package in R [54]. Age explained the highest variation in the microbiome at baseline, and hence to reduce its confounding effect, PERMANOVA models were run by controlling for age by stratifying it as blocks (strata=age). We additionally performed linear regression models with CLR transformed taxa abundances as outcomes and included potential confounders and their interaction with mental health as predictors.

## Results

### Data collection

Our study sample consisted of 369 individuals recruited from the Digbi Health research study cohort that reported anxiety/depression or sleep problems when starting Digbi Health personalized digital care intervention (Figure 1). The study subjects were enrolled on average 88.3 days (median = 64 and std = 67.7 days) by the time lost at least 2% body weight and were sent a questionnaire about their mental health and answered the questionnaire on average 1.7 days (median = 0 and std = 7.7 days) after receiving it. We obtained microbiome and genetic data for 344 and 348 individuals, respectively, 328 of whom submitted both sample types. For the baseline models of anxiety or depression, and sleep problems, all these 328 participants were studied. For the improvement models, we started with the 147, 148, and 163 individuals that reported their change in intensity for anxiety, depression, and insomnia, respectively. Most of the individuals (>95%) reported improvement or maintenance of symptoms, and only 4% (6 out of 148) for depression, 2.7% (4 out of 147) for anxiety, and 1.8% (3 out of 163) for insomnia reported higher scores at T1 compared with T0 (Figure 2 and Table S2). Due to the small sample size of individuals with worsening symptoms, we excluded them from additional analyses. Likewise, we also excluded individuals reporting improvements in 4 and 5 scale points, which were in total 8 for anxiety, 7 for depression and 6 for insomnia (Table S3). Thus, improvement models were based on 135 responses for anxiety, 135 responses for depression, and 154 responses for insomnia. We included in the analyses of the gut microbiome 178 bacterial genera and 42 functional pathways, and 23 genetic scores.

**Figure 1.**
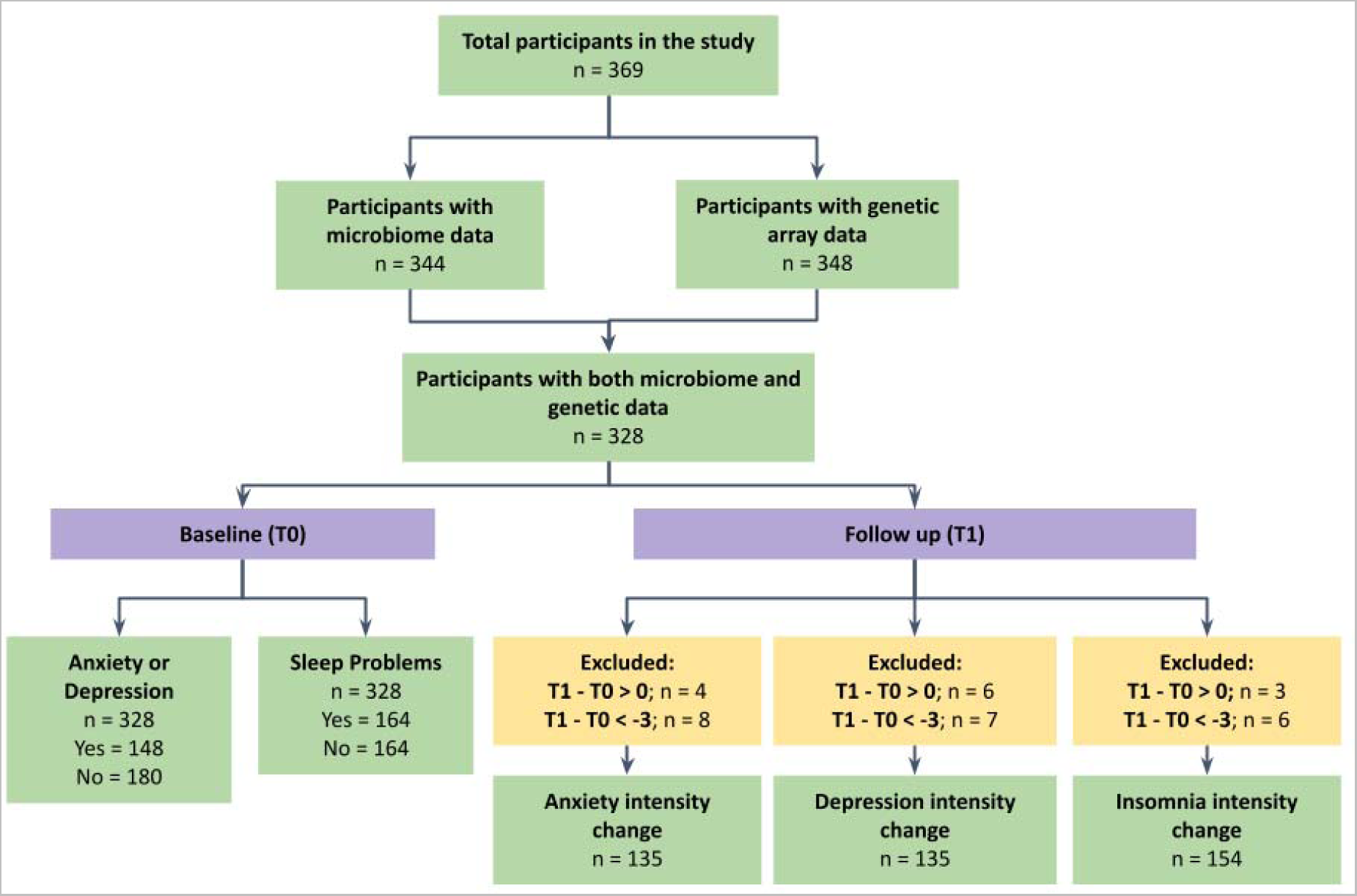
Study design and cohort sample size for each outcome. 369 subjects were recruited from the Digbi Health digital care program for whom we obtained microbiome (n = 344) and genomic SNP (n = 348) data and selected for analyses the subset of 328 subjects with both datasets (n = 328). Data were analyzed to identify gut microbiome and genetic scores associated with anxiety or depression and sleep problems at baseline (T0) and improvement in anxiety, depression, and insomnia at follow-up (T1), when subjects reached at least 2% body weight loss. For analyses at T1, we excluded individuals who had higher scores at T1 than at T0, and those improved 4 or 5 scale points, leading to sample sizes for anxiety, depression, and insomnia intensity changes of 135,135, and 154, respectively.

**Figure 2.**
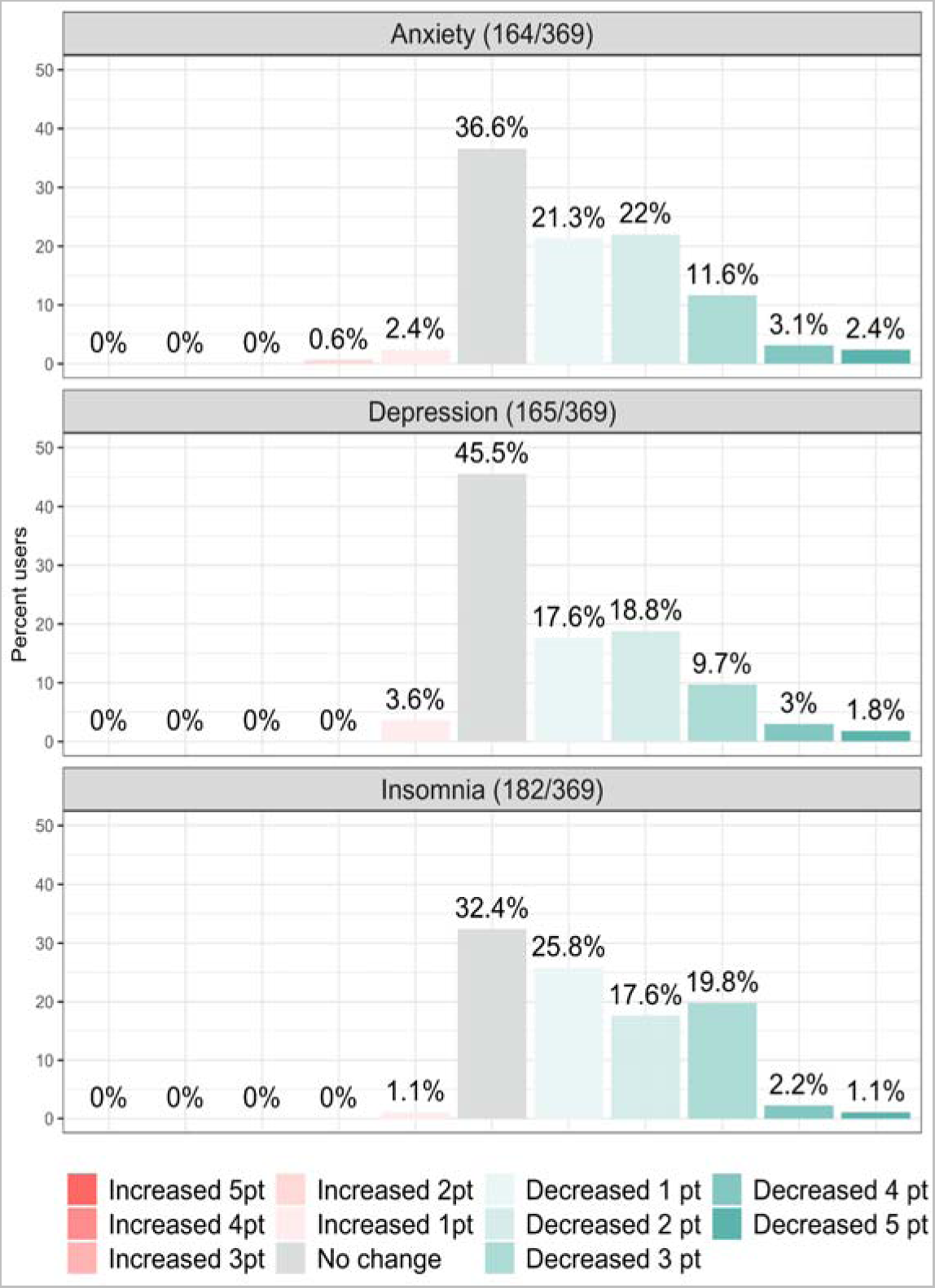
Changes in symptom intensity after the intervention. The bar plots illustrate the percent of users who self-reported symptom intensity changes in anxiety, depression, and insomnia at follow-up (T1) compared to baseline (T0), on a scale from 0 to 5.

### Cohort demographic characteristics

Table 1 summarizes the demographic variables used in the analyses performed in the study. The mean BMI of the participants was 34.6, corresponding to obese class 1 individuals and in agreement with the fact that most Digbi Health care digital program participants undergo a dietary intervention for weight loss. At the time of answering the follow-up questionnaire, most of the participants (60.4%) lost between 5 to 10% body weight. This cohort had a high prevalence of individuals with FGIDs (84%) and females (79%). 284 (86%) of the participants reported taking antidepressants (40 or 12%) or anti-anxiolytics (244 or 74%) at baseline. Table S3 provides demographic information stratified by the level of improvement in anxiety, depression, or insomnia at T1 compared with T0. Genetic ancestry analyses identified the majority of individuals as of European ancestry (43%), followed by Africans (28%), Americans (20%), east Asians (7%), and Southeast Asians (1%). We included the first five principal components from the genetic ancestry analysis as covariates in all analyses.

**Table 1.** Summary characteristics of the cohort.

| Characteristics | Baseline | intensity Improvement at T1 |  |  |
| --- | --- | --- | --- | --- |
|  | Anxiety or Depression or Sleep Problems | Anxiety | Depression | Insomnia |
| **Weight** |  |  |  |  |
| BMI at T0 | 34.6(7.3) | 35.6(8.3) | 35.6(8.3) | 34.9(7.7) |
| Normal | 4 | 2 | 2 | 2 |
| Overweight | 89 | 37 | 37 | 43 |
| Obese class 1 | 107 | 38 | 39 | 46 |
| Obese class 2 | 63 | 29 | 28 | 37 |
| Obese class 3 | 65 | 36 | 37 | 32 |
| **FGID** |  |  |  |  |
| Yes | 277 | 132 | 133 | 143 |
| No | 51 | 10 | 10 | 17 |
| **Gender** |  |  |  |  |
| Female | 258 | 119 | 122 | 133 |
| Male | 70 | 23 | 21 | 27 |
| Age | 48.7(12.2) | 46(11.7) | 45.9(11.4) | 49.3(11.4) |
| **Weight trajectory** |  |  |  |  |
| Weight gain | 2 | 1 | 1 | 1 |
| No change | 30 | 17 | 17 | 11 |
| 3-5% weight loss | 85 | 37 | 35 | 44 |
| 5-10% weight loss | 198 | 83 | 86 | 98 |
| More than 10% weight loss | 13 | 4 | 4 | 6 |
| **Medications** |  |  |  |  |
| Antidepressants | 12 | 10 | 9 | 9 |
| Antianxiolytics | 76 | 62 | 61 | 45 |
| **Total** | 328 | 142 | 143 | 160 |

### Baseline gut microbiome and genetic factors are associated with mental health improvement after dietary intervention

#### Anxiety

After the dietary and lifestyle intervention, 59% of the 135 individuals studied reported improving their anxiety symptoms, with 22%, 24%, and 13% reporting improvement in 1, 2 and 3 scale points, and 41% reporting no improvement. Three genetic scores, irritable bowel syndrome (IBS), body mass index (BMI), and obstructive sleep apnea (OSA), reached statistical significance for association with changes in anxiety (Table 2). IBS correlated with less than average improvement and BMI and OSA with greater than average improvement in anxiety intensity scores after intervention. Seven bacterial genera were statistically associated with changes in anxiety. The abundance of four, *Dorea*, *Ruminococcaceae*_*UBA1819*, *Oscillospiraceae_UCG003*, and *Eubacterium ventriosum* group, correlated with a less than average improvement, and three, *Ruminococcaceae*_*DTU089*, *Prevotella*, and *Adlercreutzia*, with a greater than average improvement in anxiety scores (Table 2). An increasing abundance of genes of the bacterial functional pathway kynurenine synthesis (MGB004) was associated with a less than average improvement in anxiety symptoms at follow-up (Table 2). Table S4, Table S5, Figure S1, and Figure S2 provide summary statistics and boxplots of the genetic scores and microbiome factors significantly associated with improvement of anxiety between T1 and T0.

**Table 2.** Variables associated with improvement of anxiety between T1 and T0. N samples correspond to the number of samples included in the analyses. In the case of microbiome analysis, it corresponds to the number of samples with an abundance greater than zero. IBS = irritable bowel syndrome, BMI = body mass index, OSA = obstructive sleep apnea and FDR = False Discovery Rate. Beta and ‘beta se’ correspond to the regression coefficient and its standard error, respectively. P-value of statistical test evaluating beta ≠0.

| Variable | N samples | beta* | beta se | p-value | FDR |
| --- | --- | --- | --- | --- | --- |
| **Genetics** |  |  |  |  |  |
| IBS | 135 | 0.248 | 0.0788 | 0.0016 | 0.015 |
| BMI | 135 | -0.218 | 0.0704 | 0.0018 | 0.015 |
| OSA | 135 | -0.232 | 0.0869 | 0.0075 | 0.04 |
| **Microbial Taxa** |  |  |  |  |  |
| *Dorea* | 118 | 0.250 | 0.081 | 0.0017 | 0.071 |
| *Ruminococcaceae_UBA1819* | 76 | 0.305 | 0.102 | 0.0028 | 0.071 |
| *Ruminococcaceae_DTU089* | 64 | -0.385 | 0.133 | 0.0039 | 0.071 |
| *Prevotella* | 51 | -0.128 | 0.045 | 0.0043 | 0.071 |
| *Oscillospiraceae_UCG003* | 65 | 0.418 | 0.148 | 0.0048 | 0.071 |
| *Eubacterium ventriosum group* | 97 | 0.238 | 0.0931 | 0.011 | 0.13 |
| *Adlercreutzia* | 77 | -0.469 | 0.190 | 0.014 | 0.15 |
| **Microbial Functions** |  |  |  |  |  |
| MGB004: Kynurenine synthesis | 135 | 0.383 | 0.131 | 0.0033 | 0.14 |

\*A value of beta > 0 indicates increasing values of the microbiome or genetic variable are associated with less than average improvement and a value of beta < 0 that the improvement is greater than average.

#### Depression

After the dietary and lifestyle intervention, 51% of the 135 individuals studied reported improving their depression symptoms, with 21%, 21%, and 9% reporting improvement in 1, 2, and 3 scale points, and 49% reporting no improvement. Table 3 summarizes the genetic and microbiome variables associated with improvement in depression intensity. Two genetic scores were directly associated with a greater than average decrease in intensity scores, OSA and AUD, and height was associated with a less than average improvement. Nine bacterial genera and four functional pathways were associated with improvement in depression intensity, eight of which were associated with less than average improvement and five with greater than average improvement (Table 3). Table S4, Table S6, Figure S3, and Figure S4 provide summary statistics and boxplots of the genetic scores and microbiome factors significantly associated with improvement of depression between T1 and T0.

**Table 3.** Variables associated with improvement of depression between T1 and T0. N samples correspond to the number of samples included in the analyses. In the case of microbiome analysis, it corresponds to the number of samples with an abundance greater than zero. OSA = obstructive sleep apnea, AUD = alcohol use disorder and FDR = False Discovery Rate. ‘Beta’ and ‘beta se’ correspond to the regression coefficient and its standard error, respectively. P-value of statistical test evaluating beta ≠ 0.

| Variable | N samples | beta* | beta se | p-value | FDR |
| --- | --- | --- | --- | --- | --- |
| **Genetics** |  |  |  |  |  |
| OSA | 135 | -0.235 | 0.088 | 0.0075 | 0.12 |
| AUD | 135 | -0.218 | 0.096 | 0.023 | 0.14 |
| Height | 135 | 0.168 | 0.076 | 0.027 | 0.14 |
| **Microbial Taxa** |  |  |  |  |  |
| *Clostridium innocuum* group | 52 | 0.183 | 0.060 | 0.0022 | 0.132 |
| *Oscillospiraceae_UCG003* | 68 | 0.283 | 0.0991 | 0.0043 | 0.132 |
| *Anaerostipes* | 132 | 0.202 | 0.0751 | 0.0071 | 0.132 |
| *Eubacterium ventriosum* group | 98 | 0.196 | 0.0746 | 0.0086 | 0.132 |
| *Lactobacillus* | 70 | -0.189 | 0.0751 | 0.011 | 0.132 |
| *Negativibacillus* | 71 | 0.279 | 0.114 | 0.014 | 0.132 |
| *Prevotella* | 52 | -0.116 | 0.0475 | 0.015 | 0.132 |
| *Oscillibacter* | 125 | 0.178 | 0.0732 | 0.015 | 0.132 |
| *Actinomyces* | 70 | -0.508 | 0.211 | 0.016 | 0.132 |
| **Microbial Functions** |  |  |  |  |  |
| MGB053: Butyrate synthesis II | 135 | 0.849 | 0.260 | 0.0011 | 0.043 |
| MGB015: p-Cresol synthesis | 135 | 0.972 | 0.341 | 0.0044 | 0.078 |
| MGB026: Nitric oxide synthesis II (nitrite reductase) | 92 | -0.167 | 0.0607 | 0.0058 | 0.078 |
| MGB027: Nitric oxide degradation I (NO dioxygenase) | 135 | -0.171 | 0.0666 | 0.0099 | 0.098 |

\*A value of beta > 0 indicates increasing values of the microbiome or genetic variable are associated with less than average improvement and a value of beta < 0 that the improvement is greater than average.

#### Insomnia

After the dietary and lifestyle intervention, 66% of the 154 individuals studied reported improving their insomnia symptoms with 29%, 18%, and 19% reporting improvement in 1, 2, and 3 scale points, and 34% reporting no improvement. Table 4 summarizes the genetic and microbiome variables associated with improvement in insomnia intensity. Two genetic scores were statistically significant, with one associated with a greater than average decrease in intensity scores, type 1 diabetes (T1D), and one associated with a less than average decrease in intensity scores, type 2 diabetes (T2D). Two bacterial genera were associated with improvement in insomnia intensity. *Butyricimonas* was associated with a less than average improvement, and *Roseburia* with a greater than average improvement. Finally, one functional pathway was associated with a greater than average improvement in insomnia, nitric oxide synthesis II (nitrite reductase). Table S4, Table S7, Figure S5, and Figure S6 provide summary statistics and boxplots of the genetic scores and microbiome factors significantly associated with improvement of insomnia between T1 and T0.

**Table 4.**
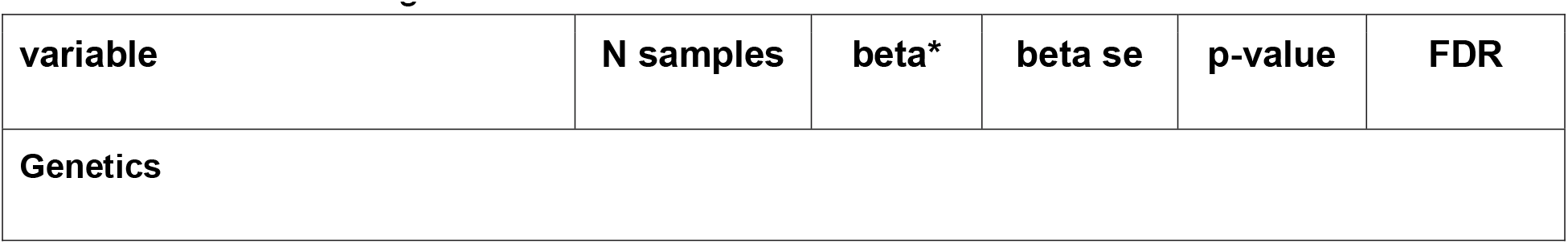
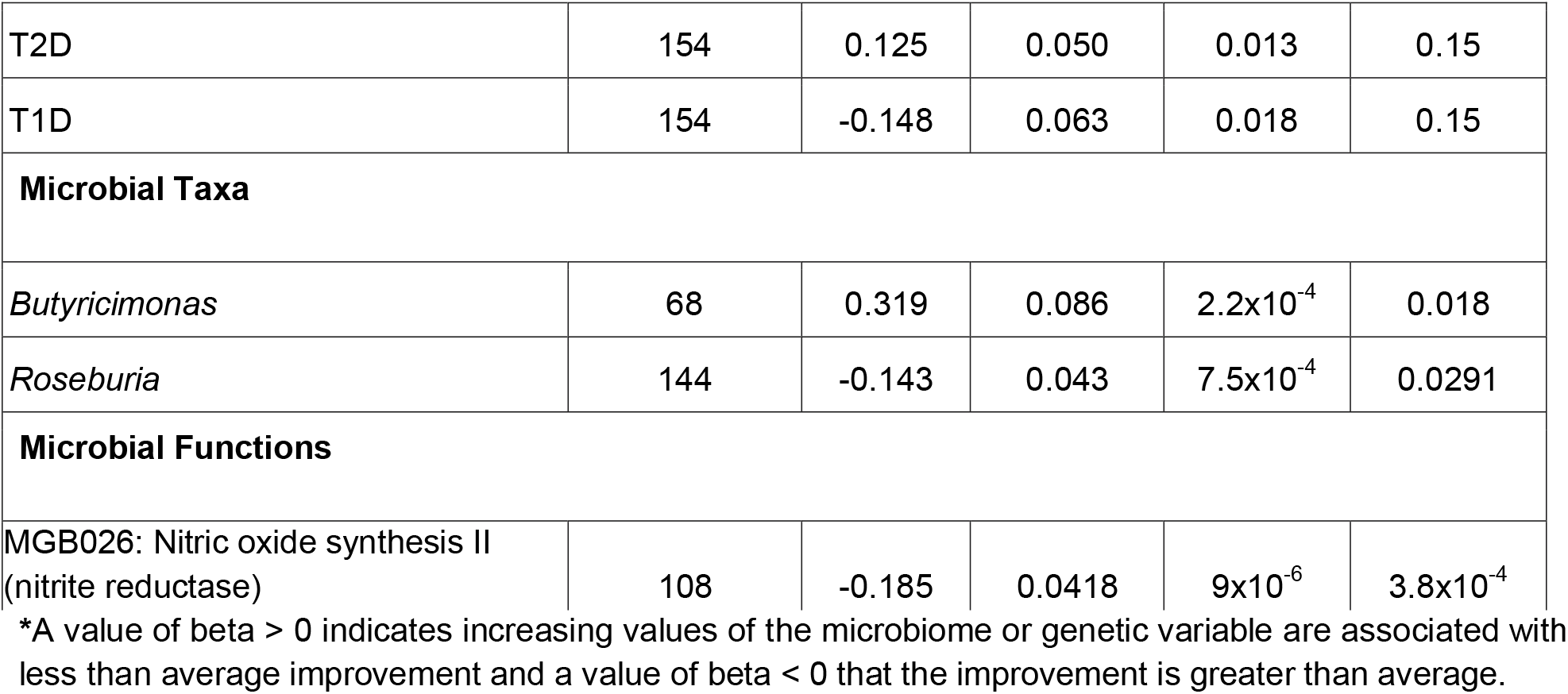
Variables associated with improvement of insomnia between T1 and T0. N samples correspond to the number of samples included in the analyses. In the case of microbiome analysis, it corresponds to the number of samples with an abundance greater than zero. T1D = type 1 diabetes, T2D = type 2 diabetes and FDR = False Discovery Rate. Beta and ‘beta se’ correspond to the regression coefficient and its standard error, respectively. P-value of statistical test evaluating beta ≠ 0.

### At baseline, psychiatric disorders’ genetic scores are associated with anxiety or depression, whereas microbial metabolic pathways associate with sleep problems

At baseline (T0), individuals that reported depression or anxiety, compared with those who did not report it, had higher mean BMI (t-test p-value = 0.022), higher prevalence of females (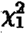 p-value = 0.021), lower mean age (t-test p-value = 2.2x10^-5^), and higher prevalence of FGIDs (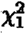 p-value = 8x10^-5^). Individuals who reported sleep problems at baseline compared with those who did not report it had a higher prevalence of FGIDs (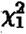 p-value = 0.021) (Table S8). We also noted significant differences in overall bacterial diversity between individuals with and without anxiety or depression at the baseline (PERMANOVA, p=0.035; Figure S7).

Additionally, we utilized logistic regression models to evaluate the association between genetic scores and microbiome components with self-reported depression or anxiety at T0 and sleep problems at T0. Two genetic scores, namely alcohol use disorder (AUD) and major depressive disorder (MDD), were statistically associated with an increased prevalence of depression or anxiety (Table 5). Two microbial functional pathways were associated with sleep problems at baseline, menaquinone synthesis (vitamin K2) I (MGB040) with increased prevalence of sleep problems at T0, and inositol degradation (MGB038) with decreased prevalence of sleep problems at T0 (Table 6). Table S9 and Table S10 provide summary statistical associations of all variables (demographics, genetic scores, microbial taxa, and functions) with anxiety or depression at baseline, and sleep problems at baseline, respectively. Table S4 provides summary statistics, and Figure S8, and Figure S9 provide boxplots of the genetic scores associated with anxiety or depression at baseline, and for the microbial functions associated with sleep problems at baseline, respectively.

**Table 5.**
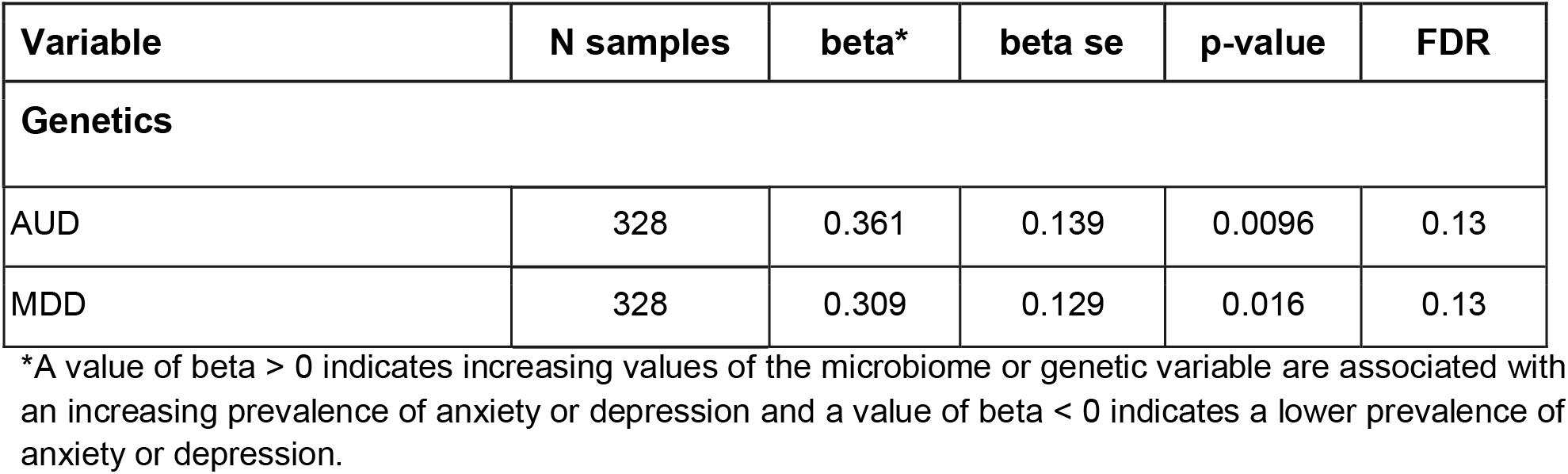
Variables associated with depression or anxiety at T0. Summary of statistical associations with depression or anxiety at baseline. N samples correspond to the number of samples included in the analyses. AUD = alcohol use disorder, MDD = major depressive disorder and FDR = False Discovery Rate. Beta and ‘beta se’ correspond to the regression coefficient and its standard error, respectively. P-value of statistical test evaluating beta ≠0.

**Table 6.** Variables associated with sleep problems at T0. Summary of statistical associations with sleep problems at baseline. N samples correspond to the number of samples included in the analyses. In the case of microbiome analysis, it corresponds to the number of samples with an abundance greater than zero. FDR = False Discovery Rate. Beta and ‘beta se’ correspond to the regression coefficient and its standard error, respectively. P-value of statistical test evaluating beta ≠ 0.

| Variable | N samples | beta* | beta se | p-value | FDR |
| --- | --- | --- | --- | --- | --- |
| **Microbial Functions** |  |  |  |  |  |
| MGB040: Menaquinone synthesis (vitamin K2) I | 328 | 0.863 | 0.298 | 0.0038 | 0.092 |
| MGB038: Inositol degradation | 328 | -0.484 | 0.170 | 0.0044 | 0.092 |

\*A value of $\beta > 0$ indicates increasing values of the microbiome or genetic variable are associated with an increasing prevalence of sleep problems and a value of $\beta < 0$ indicates a lower prevalence of sleep problems.

### Multi-omics models are better correlated with mental health improvement than demographics models alone

We compared the ability of models combining demographic (D), genetic (G), and microbiome (M) information to explain the study outcomes: anxiety or depression at baseline, sleep problems at baseline, and improvement of anxiety, depression, and insomnia from T0 to T1. Except for the improvement of insomnia symptoms, we found a trend for D+M or D+G models to explain more of the variation of the outcomes than the D model. Additionally, the D+M+G models were always better than the D models, and at least of a similar magnitude as the best D+M or D+G model (Figure 3 and Table S11).

**Figure 3.**
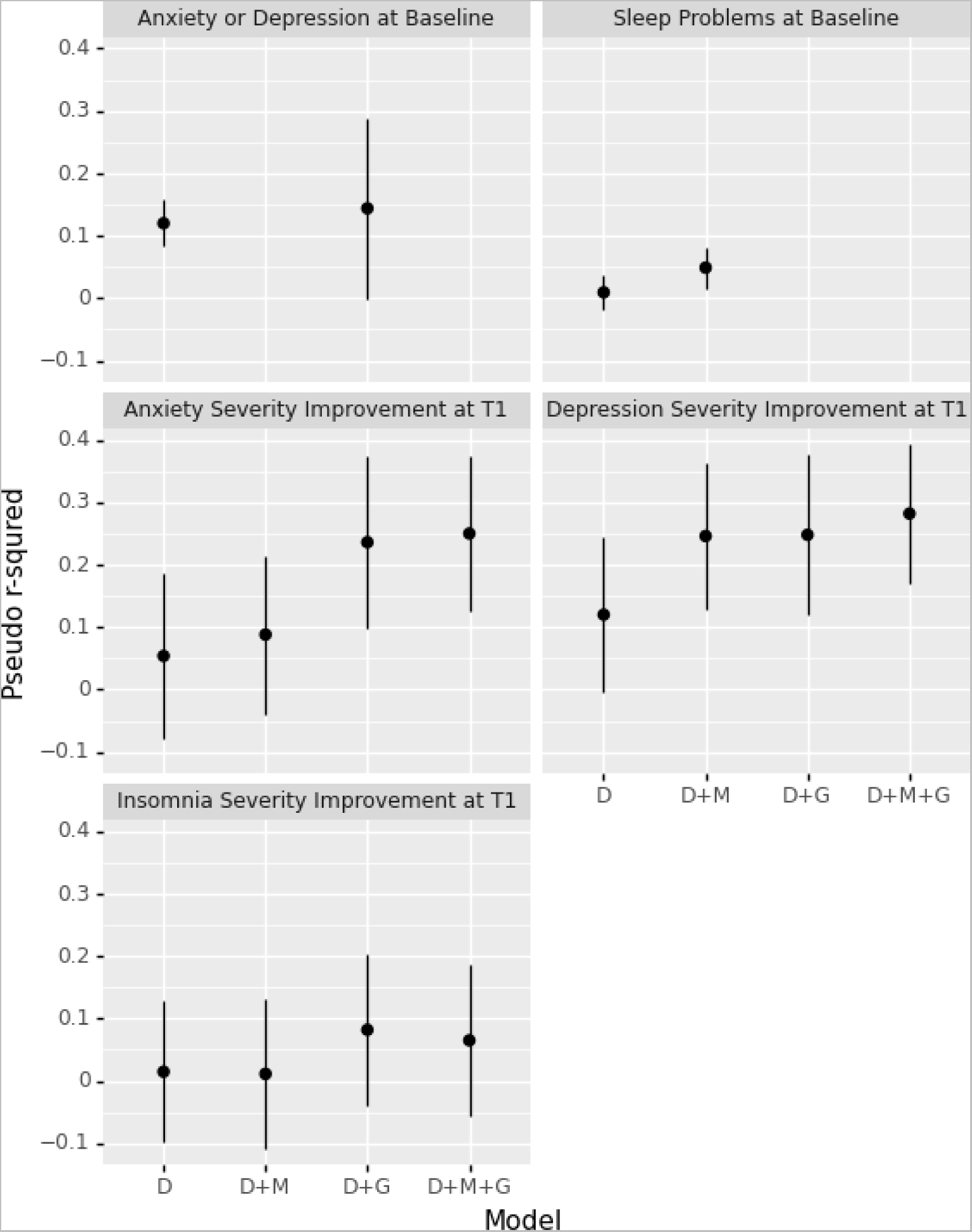
Comparison of models for all outcomes of the study based on demographic, microbiome and genetic factors. Pseudo r-square values corrected using Pratt’s method (y-axis) are shown for each model (x-axis) and outcome (different panels). The vertical line corresponds to the mean ± standard deviation obtained from 1001 bootstrap replicates. D = demographic factors only; D+M = demographic and microbiome factors; D+G = demographic and genetic factors; and D+M+G = all three sets of factors.

### Medication and recreational drug use do not confound microbiome associations with mental health

We performed sensitivity analyses to evaluate the potential confounder effect of medication, alcohol intake, and recreational drug use on the microbiome associations identified. Firstly, we performed a PERMANOVA testing the impact of the interaction between medicines and depression or anxiety at T0 or medication and sleep problems at T0 on all bacterial genera and gut-brain modules/functions (Table S12 A-D). Although we observed a marginally significant effect of anxiolytic medications (p-value = 0.04), we did not find evidence of a confounding effect of anxiolytic or antidepressant drugs on either anxiety or depression at T0 (p-value = 0.517 and 0.762, respectively) or sleep problems at T0 (p-value = 0.82 and 0.62, respectively). Similarly, we did not find any confounding effect of medications when the analysis was repeated with a subset of microbial markers (both bacterial genera and functions) that were found to be significantly associated with the mental health status at baseline and change in intensity of a particular outcome at T1 (Table S12 E and F). Lastly, we performed the same analyses for each genus and pathway separately. We found no confounding effect of medication, alcohol intake, or recreational drug use on the bacterial genera and pathways identified as significantly associated with the outcomes. The only marginal association, which did not pass multiple testing corrections, was MGB038 (inositol degradation pathway) and alcohol consumption associated with sleep problems at T0 (p-value = 0.017).

We also noted that demographic factors such as age (controlled for age by stratifying it as blocks in all PERMANOVA models), gender, and BMI explained most of the variation in the gut microbiome at baseline. We accounted for these factors as covariates in all the statistical models, including the multivariate models. We observed no significant confounding effect of these factors on the associations of the microbiome with mental health outcomes (Table S12). Furthermore, we also did not find any significant association between alcohol consumption or use of recreational drugs and baseline gut microbiome (Table S12).

## Discussion

We identified demographic, genetic, and gut microbiome factors that correlate with the mental health status at baseline and future improvement in mental health outcomes, particularly depression, anxiety, and insomnia. Overall, study participants lost on average 5.4% body weight during the study, and more than 95% reported having an improvement in at least one mental health outcome (Figure 2 and Table S2).

Our study evaluated the association between genetic scores and microbiome factors with the improvement in depression, anxiety, and insomnia after a dietary and lifestyle intervention. Therefore, this study assessed a gene and microbiome by environment interaction where the individuals responded and improved differentially depending on their genetic and baseline microbiome profile. Previous research has focused on identifying genetic and microbiome correlations and signatures with disease diagnosis or symptoms severity with genetic studies robustly listing many genetic factors and microbiome studies beginning to provide compelling evidence of association with multiple mental health conditions (for instance, see [5, 15, 55, 56]). Currently, it is unknown whether these genetic or microbiome factors are also associated with improvement after treatment, primarily since different interventions may act through different physiological mechanisms. To address this question, we compared the association between genetic and microbiome factors and anxiety or depression, and sleep problems at baseline versus anxiety, depression and insomnia intensity change at follow-up time (T1). We found the effect size of the association between the outcomes at the two different time points is correlated but weakly so, with microbial functional pathways having the largest r-squared values, for instance, 0.38 for the association between anxiety or depression at baseline with depression improvement at T1 (Figure S10, Figure S11, and Figure S12). Nonetheless, several of the associations we identified with improvement in anxiety, depression, or insomnia had been previously associated with the absence of these conditions. Thus, the strategy of using association with diagnosis to identify signatures with validity to explain treatment response warrants further research as more robust genetic, and gut microbiome associations with different diagnoses are being reported.

This study identified 8 genetic scores, 15 microbiome genera, and 7 functional pathways associated with improvement in anxiety, depression, insomnia, or anxiety/depression and sleep problems at baseline. We noted an association between a higher abundance of kynurenine synthesis (MGB004) and a less than average improvement in anxiety intensity. Kynurenine is a catabolic product of the tryptophan-kynurenine metabolism and it is further metabolized into kynurenic acid or quinolinic acid. Previous studies have reported higher levels of plasma kynurenine to be associated with anxiety [57] and higher plasma levels of quinolinic acid associated with depression [58, 59]. The gut microbial pathway involved in p-cresol synthesis (MGB015) was strongly associated with a less than average improvement in depression intensity. Although previous reports suggest the role of this gut microbial metabolite in autism [60], its role in depression has not been clearly identified.

Interestingly, nitric oxide synthesis II (MGB026: nitrite reductase) was significantly associated with a greater than average improvement in depression and insomnia intensity. In contrast, the nitric oxide degradation I pathway (MGB027: nitric oxide dioxygenase) was associated with a greater than average improvement in depression intensity only. The association between nitric oxide degradation I (NO dioxygenase) (MGB027) and improvement in depression intensity is of great interest. Increased NO levels have been found in the plasma of MDD patients (see, for instance, [61]). Antidepressants and anxiolytics have been shown to induce inhibition of NO synthesis (see [62] and references therein). Therefore, increased NO degradation by the gut microbiome may mimic the effects of pharmacological treatments. Research studies provide physiological context for the association between NO and sleep and anxiety and mental health more generally reviewed by [62, 63]. Contrastingly, the research literature also suggests that higher NO levels may be beneficial for mental health due to its role in neuronal plasticity, inflammation, and oxidative stress [64]. Given its intrinsic properties enabling rapid diffusion and activation of signaling cascades with functions in multiple physiological contexts, it is not surprising that the mechanistic link between NO degradation or synthesis by the gut microbiome and human behaviors is not well understood yet.

The association between the increasing abundance of butyrate synthesis II (MGB053) and a less than average improvement in depression is in the opposite direction of the generally reported relationship between short-chain fatty acids (SCFAs) and mental health [62, 65]. These previous reports link lower butyrate with poorer mental health before intervention. However, the association found in this study relates to the effect of the intervention, and thus reflects that a high abundance of butyrate synthesis genes at baseline is associated with improvement but less so than the average. This could be explained because the intervention increases dietary fiber, which is known to increase the relative abundance of butyrate-producing microbes and those producing other SCFAs. Therefore, having a high baseline abundance of butyrate synthesis genes may limit the beneficial effect an individual can attain during the intervention. We also note that the literature reports that the beneficial or detrimental effects of higher butyrate synthesis by the gut microbiome may depend on the context, such as the section of the intestine where the butyrate-producing microbes are inhabiting [66].

Previous research has pointed to the association between depression and *Oscillospiraceae_UCG003* [67], *Eubacterium ventriosum* group [68, 69], *Lactobacillus* [70], *Prevotella* [71], and anxiety with *Ruminococcaceae*_*UBA1819* [72] and *Ruminococcaceae*_*DTU089* [73] and the direction of the association was concordant with previous reports. *Butyricimonas* and *Roseburia* were associated with improvement in insomnia, replicating previously reported associations [74]. Interestingly, several genera were systematically associated with anxiety or depression at baseline and with improvement in multiple outcomes. For instance, the *Eubacterium ventriosum* group was significantly associated with improvement in anxiety and depression with the same direction of effect (beta = 0.24 and 0.20). Likewise, *Prevotella* was associated with improvement in anxiety and depression with the same direction of effect (beta = -0.13 and -0.12) and nominally associated with anxiety or depression at baseline (beta = -0.3 and p-value = 0.0028) (Table S9C). *Oscillospiraceae_UCG003* was associated with improvement on depression (beta = 0.28) and reached a nominal association with improvement on anxiety (beta = 0.42 and p-value = 0.0048) (Table S5C). In fact, there were no genera associated (with p-values < 0.05) with multiple outcomes with an inconsistent direction of effect.

The association between the IBS genetic score and a less than average improvement in anxiety suggests that individuals at higher inherited risk for IBS improve their anxiety symptoms less than those without the risk after our digital therapeutics intervention. Cameron et al. [21] had already reported a positive correlation between genetic risk for IBS and anxiety, and Eijsbouts et al. (2021) suggest a common basis for anxiety and IBS independent of their comorbidity. Our results can be interpreted as evidence that this shared etiology may also have implications for therapeutic response. The association between BMI and OSA genetic scores with a greater than average improvement in anxiety may be explained based on the direct relationship between obesity, and the occurrence of OSA [75] and the fact that sleep disturbances and poor sleep are partially caused by OSA, and OSA is associated with higher anxiety symptoms [76, 77]. Therefore, it is plausible that weight loss would lead to a reduction in OSA and anxiety. In line with his hypothesis, examining the ratio of the linear regression coefficients for improvement of anxiety for different weight loss groups shows that subjects who did not lose or gain body weight (No change group) improved 17% less (No change/3-5% weight loss = 0.6163/-3.6632 = -0.168, p-value = 0.003), and those that gained weight (Weight gain group) improved 62% less (Weight Gain/3-5% weight loss = 2.273/-3.6632 = -0.620, p-value = 0.025) than those that lost between 3-5% percent body weight (Table S5). Other weight loss groups did not differ significantly in improvement compared to the 3-5% weight loss group.

Improvement in depression was associated with the AUD genetic score, with a higher genetic score implying a more significant improvement in self-reported depression. Genetic scores for alcohol use are positively correlated with the amount of alcohol consumed. Avoiding alcohol consumption is strongly recommended as part of the digital therapeutics intervention, and removing alcohol from the diet would lead to improvement in mental health.

Epidemiological studies have shown significant comorbidity between insomnia and type 1 and type 2 diabetes [78, 79], and there exists evidence showing that weight loss is associated with improvement in sleep and insomnia [80]. In line with our results associating the T1D genetic score with a greater than average improvement of insomnia, previous research has linked autoimmune disease and T1D in particular with increased risk of insomnia. Interestingly, we also found the T2D genetic score associated with a less than average improvement in insomnia. In addition to their co-occurrence, there exists evidence of a genetic [81] and a causal link between T2D and insomnia as supported by two out of three genetic scores identified by the MR-Base database by Hemani *et al.*, with the only discordance occurring on an automatically calculated genetic score [82]. The metabolic nature of T2D, compared with T1D, is more prone to improvement under the implemented dietary intervention, which is focused, among other objectives, to reduce insulin resistance and diabetes severity and risk, which could explain why subjects with higher genetic risk for T2D may improve less than average on their insomnia.

Subjects’ reported anxiety or depression at baseline was associated with the genetic scores of two psychiatric disorders, namely alcohol use disorder (AUD) and major depressive disorder (MDD), with a higher risk of self-reporting anxiety or depression with increasing genetic score values. These two associations align well with known genetic correlations of AUD and MDD with mood and anxiety disorders [83, 84]. We also identified an association between gut microbiome menaquinone synthesis (vitamin K2) I (MGB040) and a higher prevalence of sleep problems at baseline. Despite the paucity of evidence documenting this relationship, previous research provides context for our findings [85–88] and warrants additional investigation. Similarly, we noted a significant association of microbial inositol degradation (MGB038) with a lower prevalence of sleep-related issues at baseline. A few early reports suggest an association of frontal cortex myo-inositol concentration with sleep and depression [89]. Previous evidence suggests a possible mechanism through the relationship between Inositol and serotonin receptors [90, 91]. Although there exists no direct evidence of the relationship between gut microbial-based inositol degradation and sleep improvement, the higher abundance of bacterial pathways to catabolize inositol may be due to higher inositol levels in the body. It would be worthwhile to validate these findings using targeted multi-omics approaches.

This study has some limitations that are important to note. Firstly, the findings from this study are derived from a weight loss cohort and thus may be only reflective of the population with mental health that is overweight or obese or that may benefit from weight loss. Secondly, the inclusion criteria in this study did not consider factors known to influence the microbiome composition (e.g., probiotic) or other comorbidities (pain, skin conditions, musculoskeletal, cholesterol, diabetes, hypothyroidism, or hypertension) that may confound the results presented. Thirdly, the survey instrument utilized was an ad-hoc questionnaire that asked participants to rate their presence/absence at baseline of anxiety or depression and insomnia and their symptom intensity on a scale of 1 to 5 for anxiety, depression, and insomnia, and was not a validated clinical instrument. Additionally, the survey was performed retrospectively for the baseline (or T0) time point after subjects successfully achieved 2% or more body weight loss, so the data may contain recall bias. Furthermore, our questionnaire did not measure environmental health, social determinants of health, or any other situational factors that could contribute to mental health changes during the study period. Despite being a retrospective and self-reported measure of depression or anxiety, we believe our phenotypic assessment has validity. Self-reported depression is a valid construct to identify genetic factors associated with MDD and yields valuable findings. However, it has to be taken into consideration that cases will also include a broader set of psychopathologies, including anxiety and neuroticism [56, 83]. In addition, we observe a high concordance, 81% (74 out of 91), between the self-reported measure of depression or anxiety and the self-reported use of medication for either condition, in line with published research [92]. Fourthly, we used statistical inference algorithms to obtain information on the relative level of bacterial functional pathways, an indirect measure of the abundance of those genes that do not directly relate to the enzymatic activity or molecule levels. We believe our findings are relevant for the large population of individuals who are affected by mental health issues and obesity, and we found that they are also congruent with previous reports. However, they warrant validation via replication on independent samples and follow-up studies using metabolomics, longitudinal microbiome sampling, and other assays to understand the associations in more depth.

## Conclusions

The data and evidence gathered in this study support the notion that weight loss is associated with improvement in mental health and that there exists heterogeneity in the level of improvement among subjects. Genetic and gut microbiome factors contribute to and mediate individuals’ improvement in mental health after dietary intervention with relative importance equal to or greater than that of demographic characteristics on their own. Our results provide evidence of the value of genetic and microbiome factors to explain future mental health improvement through a digital therapeutics intervention and will help guide further personalization of interventions.

## Author statements

### Author contributions

IP and SK: formal analysis, methodology, software, visualization, writing – original draft, and writing – review and editing. BJ, DM, TU, KM: formal analysis software. SS, SS-R, CI, CR-S: formal analysis, methodology. PD: conceptualization, and writing – review and editing. DA: conceptualization, writing – original draft, and writing – review and editing. RS: conceptualization, funding acquisition, and writing – review and editing. All authors contributed to the article and approved the submitted version.

## Data Availability

The microbiome sequence data used in this study were submitted to NCBI SRA under Bioproject accession number PRJNA821674.

https://www.ncbi.nlm.nih.gov/bioproject/PRJNA821674

## Acknowledgments

We are grateful to subjects participating in Digbi Health research studies without whom it would not have been possible to perform this research.

## Supplementary Table Legends

**Table S1. Genetic scores used in the study.** For each genetic score used we provide its abbreviation as used on the main text, the number of genetic variants, reference to the original scientific study, and GWAS Catalog identifier when available.

**Table S2. Demographic data of participants and survey information used in the study.**

**Table S3. Summary characteristics of the cohorts studied for the improvement of anxiety, depression, and insomnia.** We provide mean and standard deviation (i.e. mean(std)) for BMI at T0 and Age, and counts for other variables included in all models. P-values were calculated with a t-test to compare means and a chi-squared test for count tables.

**Table S4. Summary statistics of genetic and microbiome factors significantly associated with the study outcomes.** For each of the outcomes, we provide the mean and standard deviation of the genetic scores and microbiome taxa and pathways identified as statistically significant.

**Table S5. Statistical associations with improvement in anxiety. A.** Associations with demographic variables. **B.** Associations with genetic scores. **C.** Associations with microbiome genera. **D.** Associations with microbiome functional pathways.

**Table S6. Statistical associations with improvement in depression. A.** Associations with demographic variables. **B.** Associations with genetic scores. **C.** Associations with microbiome genera. **D.** Associations with microbiome functional pathways.

**Table S7. Statistical associations with improvement in insomnia. A.** Associations with demographic variables. **B.** Associations with genetic scores. **C.** Associations with microbiome genera. **D.** Associations with microbiome functional pathways.

**Table S8. Summary characteristics of the cohorts studied at baseline.** We provide mean and standard deviation (i.e. mean(std)) for BMI at T0 and Age, and counts for other variables included in all models. P-values were calculated with a t-test to compare means and a chi-squared test for count tables.

**Table S9. Statistical associations with anxiety or depression at baseline. A.** Associations with demographic variables. **B.** Associations with genetic scores. **C.** Associations with microbiome genera. **D.** Associations with microbiome functional pathways.

**Table S10. Statistical associations with sleep problems at baseline. A.** Associations with demographic variables. **B.** Associations with genetic scores. **C.** Associations with microbiome genera. **D.** Associations with microbiome functional pathways.

**Table S11. Model fit of combined models of demographics, microbiome, and genetic predictors.** For each outcome, we performed a bootstrap analysis with 1001 replications of the models including demographic predictors (D), demographic and microbiome predictors (D+M), demographic and genetic predictors (D+G), and all three sets of predictors (D+M+G). For microbiome and genetic predictors, we included in the models variables identified on univariate analyses with an FDR ≤ 0.15. Model fit is reported as the median, mean and standard deviation of the Cox-Snell pseudo r-squared obtained from 1001 bootstrap replicates.

**Table S12. Effect of medications on the gut microbiome.** Statistical associations of self-reported intake of anxiolytics and depression medications on the gut microbiome. The potential confounding effect of the medication on the results of the study is indicated by the association of the interaction between medication and the study outcomes. Panels A through F show multivariate analyses based on Bray-Curtis dissimilarity to test the influence of covariates on the gut microbiome and anxiety or depression at baseline, or sleep issues at baseline. PERMANOVAs were performed with 999 bootstrap iterations. **A.** PERMANOVA using bacterial taxa at the genus level (genera: n=178) to test the influence of covariates on the gut microbiome and anxiety or depression at baseline. **B.** PERMANOVA using bacterial taxa at the genus level (genera: n=178) to test the influence of covariates on the gut microbiome and sleep issues at baseline. **C.** PERMANOVA using bacterial functions (gut-brain modules: n=42) to test the influence of covariates on the gut microbiome and anxiety or depression at baseline. **D.** PERMANOVA using bacterial functions (gut-brain modules: n=42) to test the influence of covariates on the gut microbiome and sleep issues at baseline. **E.** PERMANOVA using only bacterial genera and functions with significant association from the regression models (genera: n=15, and gut-brain modules: n=7) to test the influence of covariates on the gut microbiome and anxiety or depression at baseline. **F.** PERMANOVA using only bacterial genera and functions with significant association from the regression models (genera: n=15, and gut-brain modules: n=7) to test the influence of covariates on the gut microbiome and sleep issues at baseline. **G.** Univariate analysis to test the influence of covariates on the gut microbiome and outcomes of interest. The table describes results from linear regression analyses using bacterial CLR transformed abundances and as covariates gender, age, BMI at T0, and weight loss group (weight Gain > 3%, No Change, Weight loss 3-5%, Weight loss 5-10% and Weight loss +10%) using only bacterial genera and functions with significant associations to four outcomes, namely anxiety intensity change, depression intensity change, insomnia intensity change, and sleep problems at T0. For each outcome and associated microbiome factor, we report beta and beta se corresponding to the regression coefficients and their standard error, respectively. z and P-value of statistical test evaluating beta ≠ 0. ’0.025’ and ’0.975’ correspond to the confidence intervals of the beta. anx_change, dep_change, and ins_change are changes in anxiety, depression, and insomnia from T0 to T1.

---

**Figure S1.**
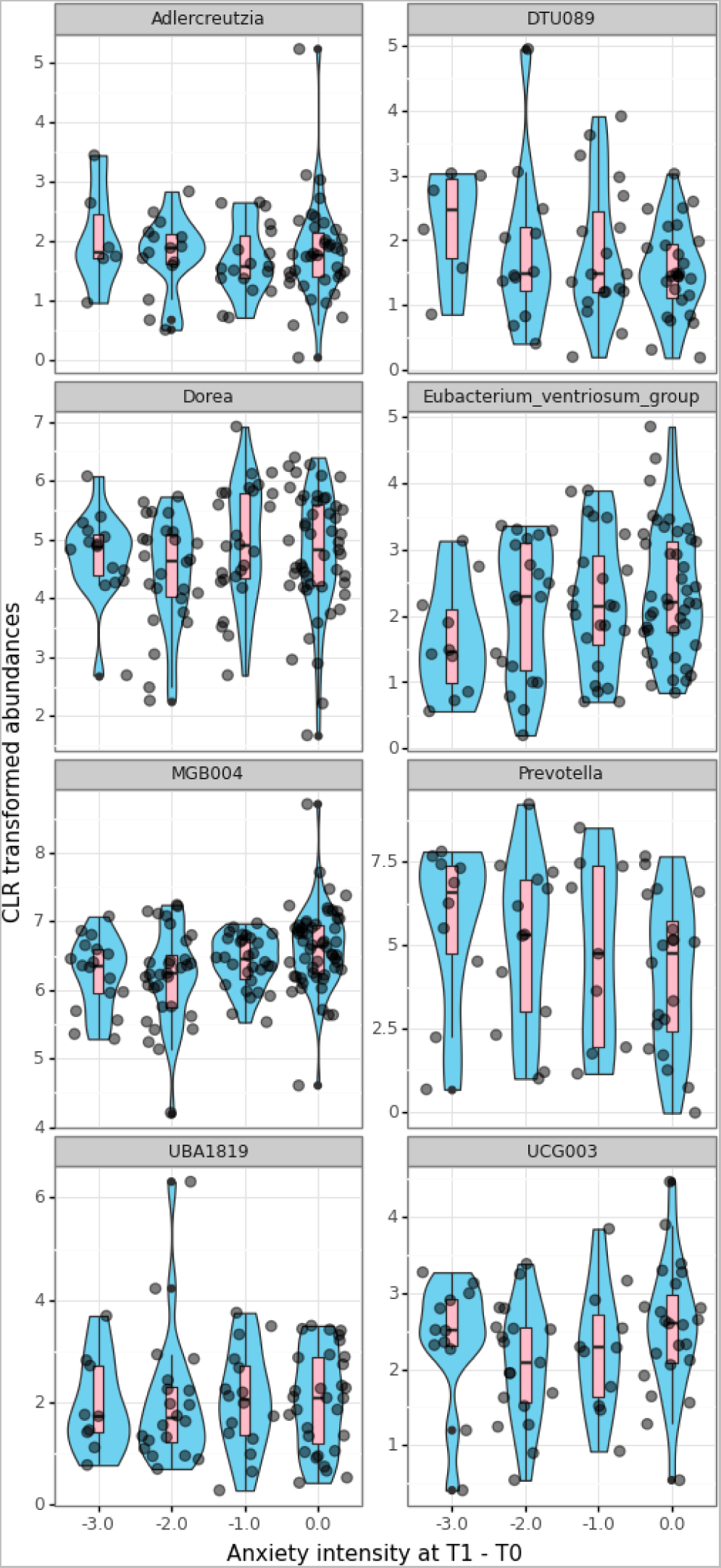
Boxplot of microbiome features associated with improvement on anxiety. Each panel provides violin plots (sky blue fill), box plots (pink fill), and horizontally jittered points presenting CLR transformed abundances of bacterial genera and functional pathways (y-axis) for each level of improvement of intensity on the outcome measured at T1 - T0 (x-axis). Panel titles display the name of the respective microbiome feature.

**Figure S2.**
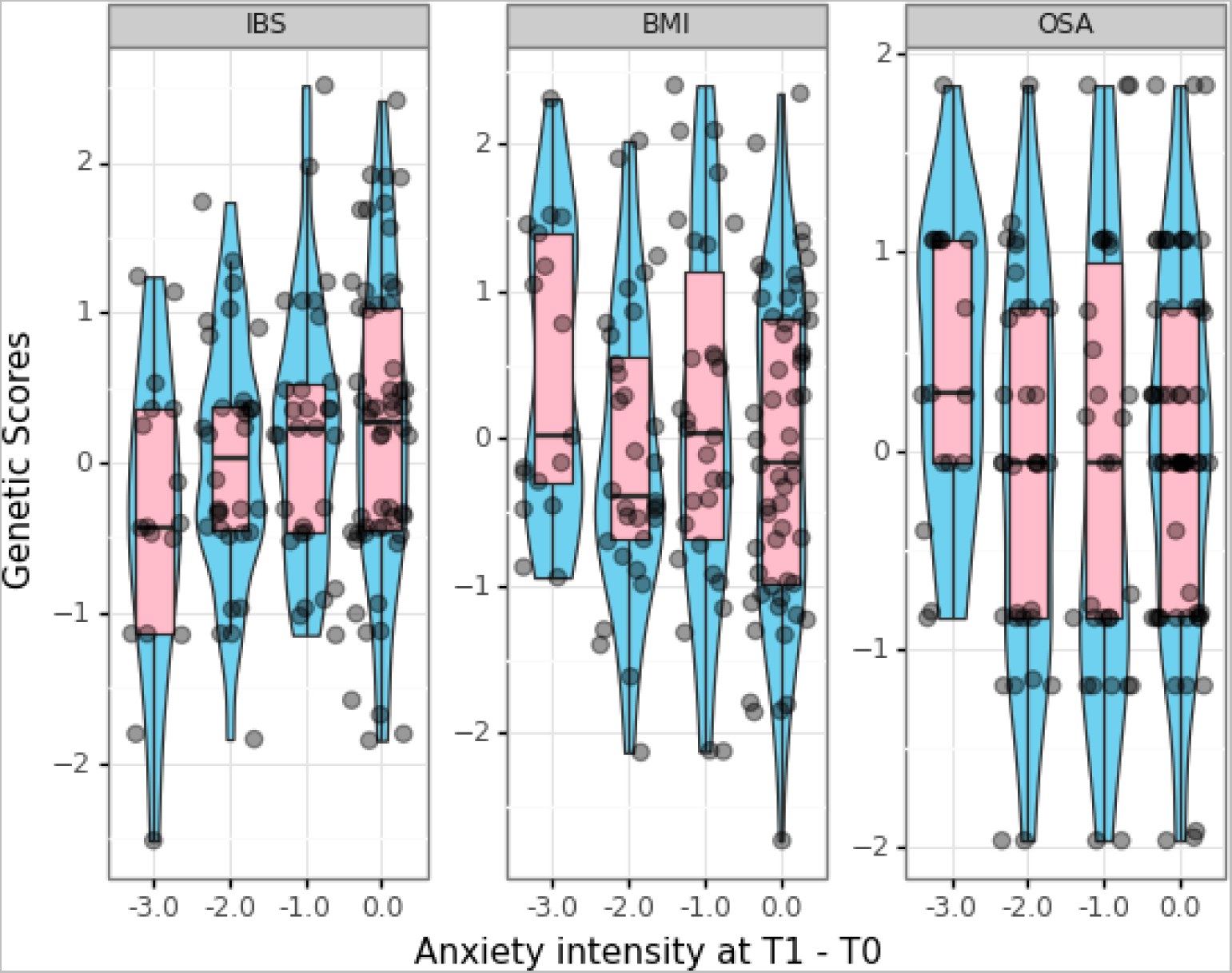
Boxplot of genetic scores associated with improvement on anxiety. Each panel provides violin plots (sky blue fill), box plots (pink fill), and horizontally jittered points presenting genetic score values (y-axis) for each level of improvement of intensity on the outcome measured at T1 - T0 (x-axis). Panel titles display the name of the respective genetic score.

**Figure S3.**
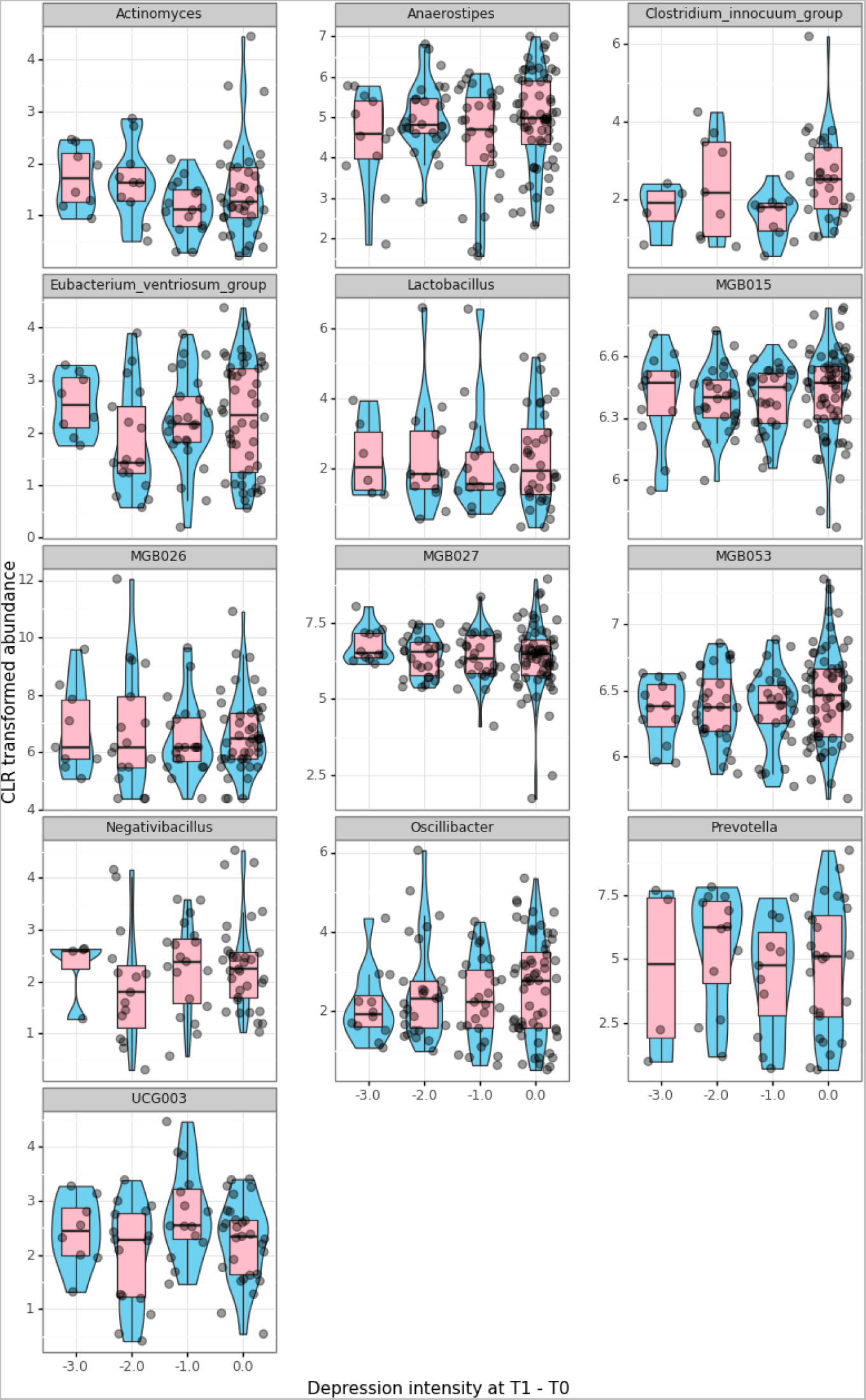
Boxplot of microbiome features associated with improvement on depression. Each panel provides violin plots (sky blue fill), box plots (pink fill), and horizontally jittered points presenting CLR transformed abundances of bacterial genera and functional pathways (y-axis) for each level of improvement of intensity on the outcome measured at T1 - T0 (x-axis). Panel titles display the name of the respective microbiome feature.

**Figure S4.**
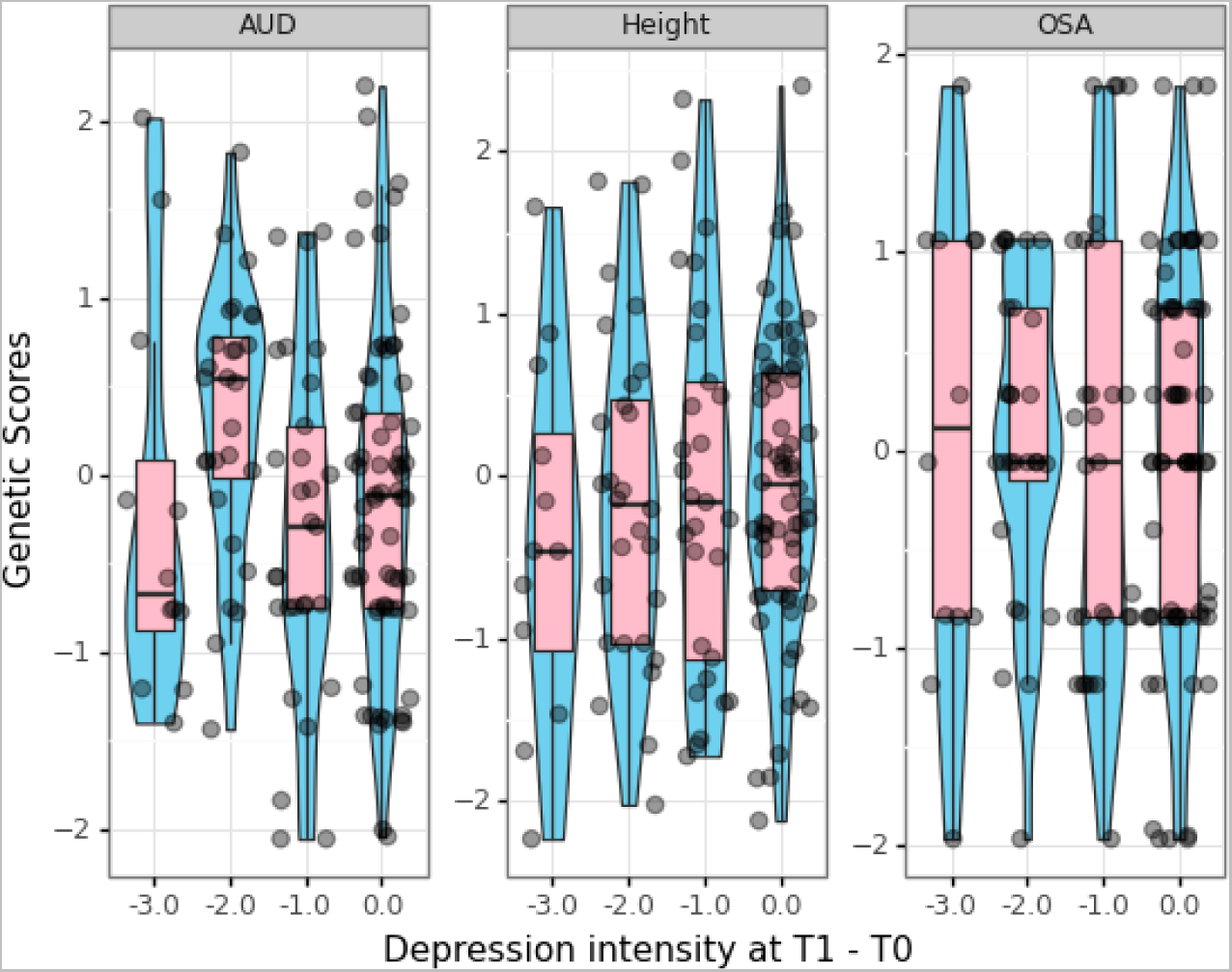
Boxplot of genetic scores associated with improvement on depression. Each panel provides violin plots (sky blue fill), box plots (pink fill), and horizontally jittered points presenting genetic score values (y-axis) for each level of improvement of intensity on the outcome measured at T1 - T0 (x-axis). Panel titles display the name of the respective genetic score.

**Figure S5.**
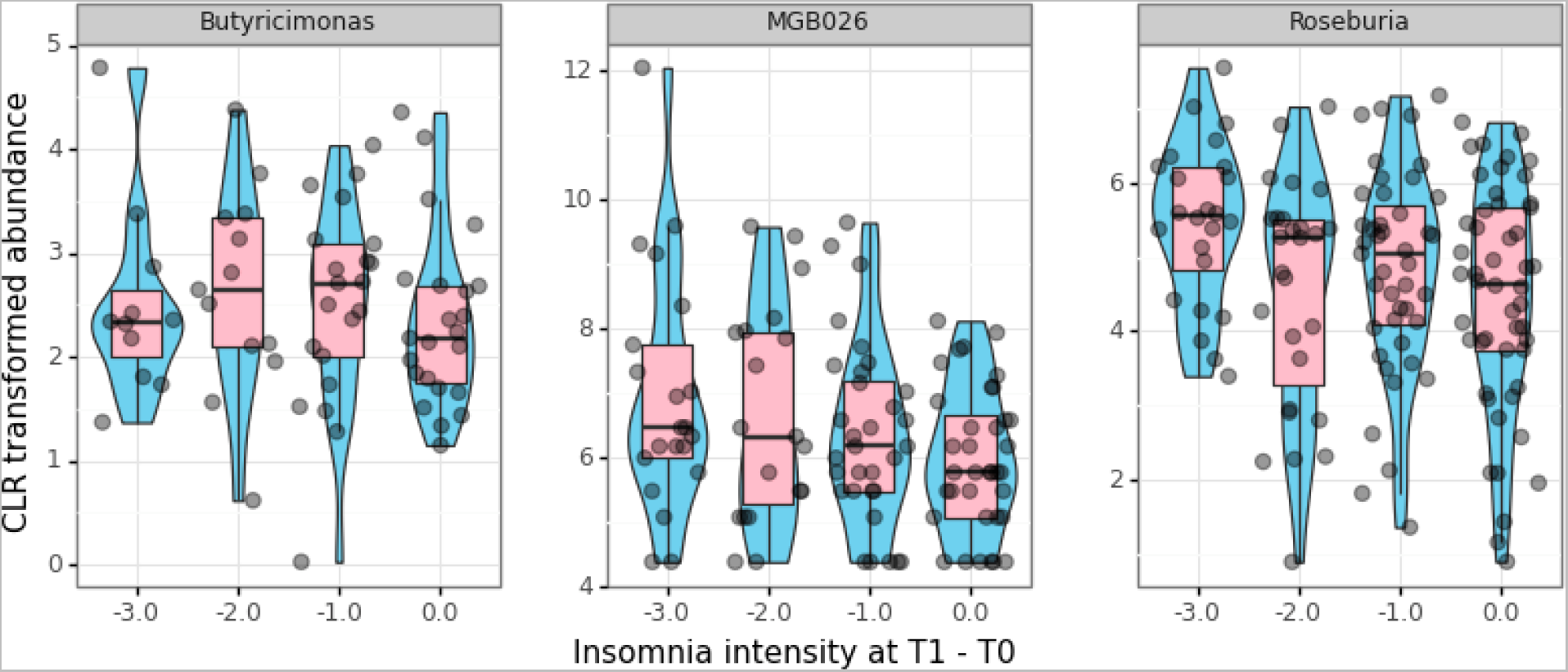
Boxplot of microbiome features associated with improvement on insomnia. Each panel provides violin plots (sky blue fill), box plots (pink fill), and horizontally jittered points presenting CLR transformed abundances of bacterial genera and functional pathways (y-axis) for each level of improvement of intensity on the outcome measured at T1 - T0 (x-axis). Panel titles display the name of the respective microbiome feature.

**Figure S6.**
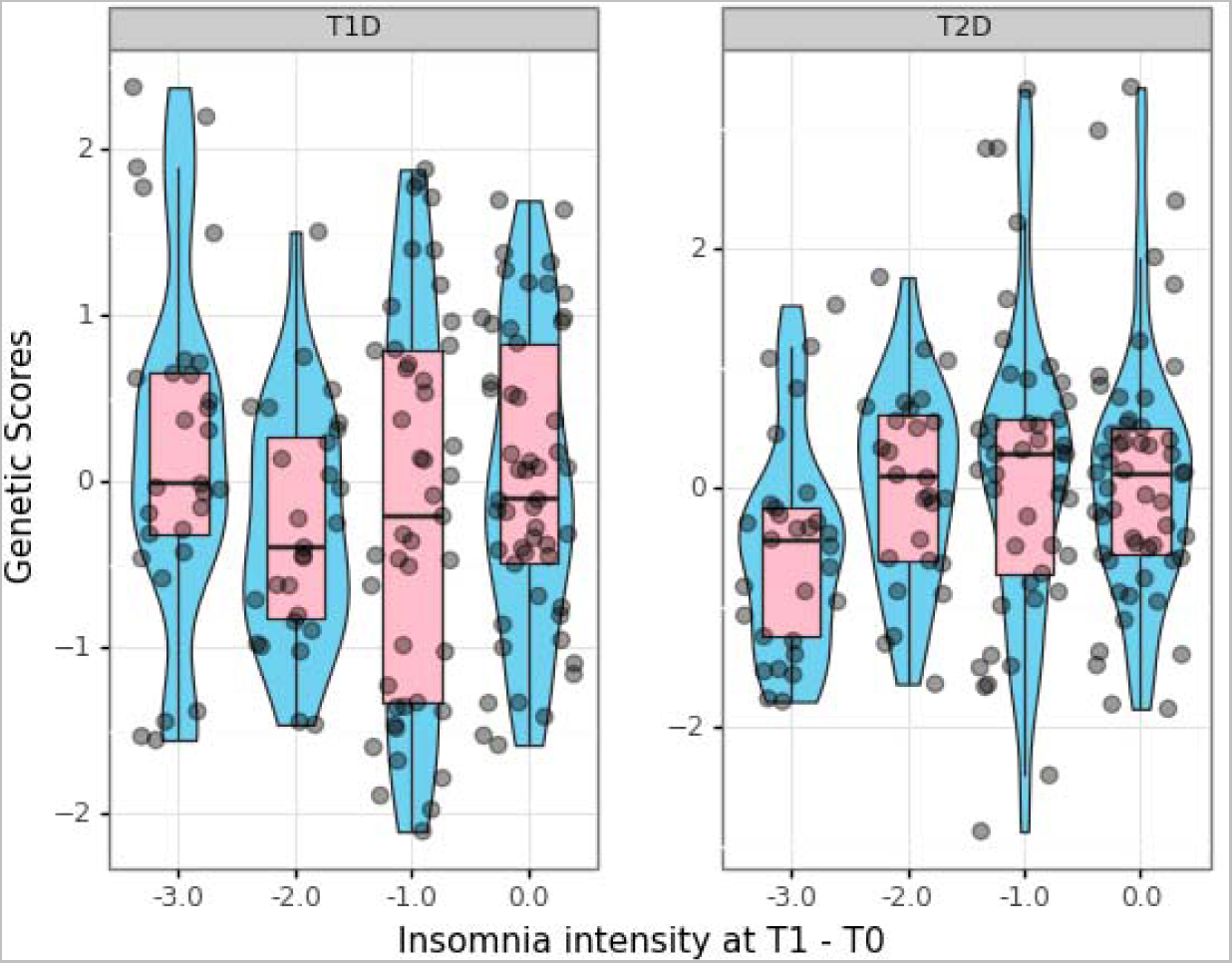
Boxplot of genetic scores associated with improvement on insomnia. Each panel provides violin plots (sky blue fill), box plots (pink fill), and horizontally jittered points presenting genetic score values (y-axis) for each level of improvement of intensity on the outcome measured at T1 - T0 (x-axis). Panel titles display the name of the respective genetic score.

**Figure S7.**
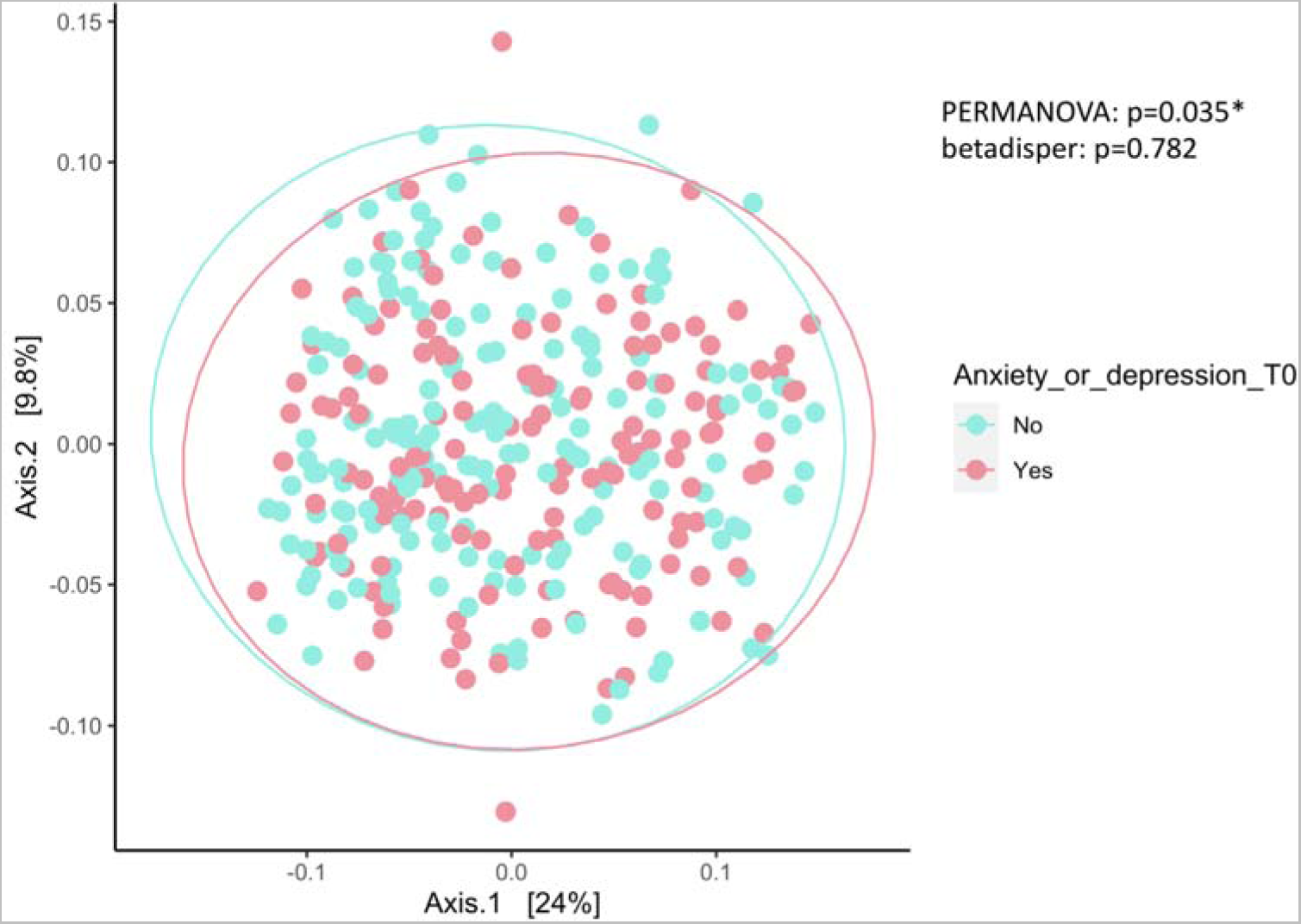
Community-level comparison of the gut microbiome of 328 individuals with and without reported anxiety or depression at baseline. The figure illustrates a PCoA plot using Bray-Curtis dissimilarity at the genus level. P-values from: PERMANOVA (with strata=age) and beta dispersion test (homogeneity of variances). Ellipses around each group represent a multivariate t-distribution and are based on the variance observed among each group of samples.

**Figure S8.**
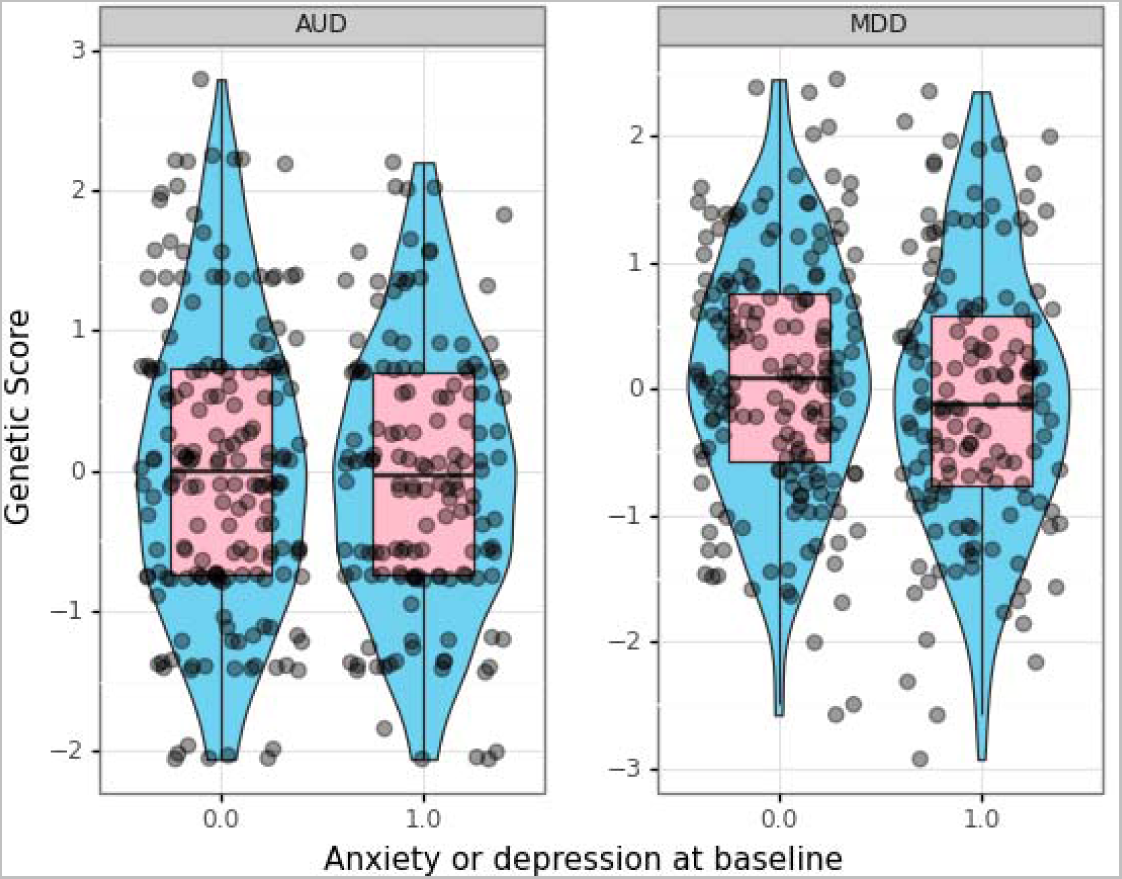
Boxplot of genetic scores associated with anxiety or depression at baseline. Each panel provides violin plots (sky blue fill), box plots (pink fill), and horizontally jittered points presenting genetic score values (y-axis) for each level of the outcome measuring the reported anxiety or depression at baseline: yes = 1 and no = 0 (x-axis). Panel titles display the name of the respective genetic score.

**Figure S9.**
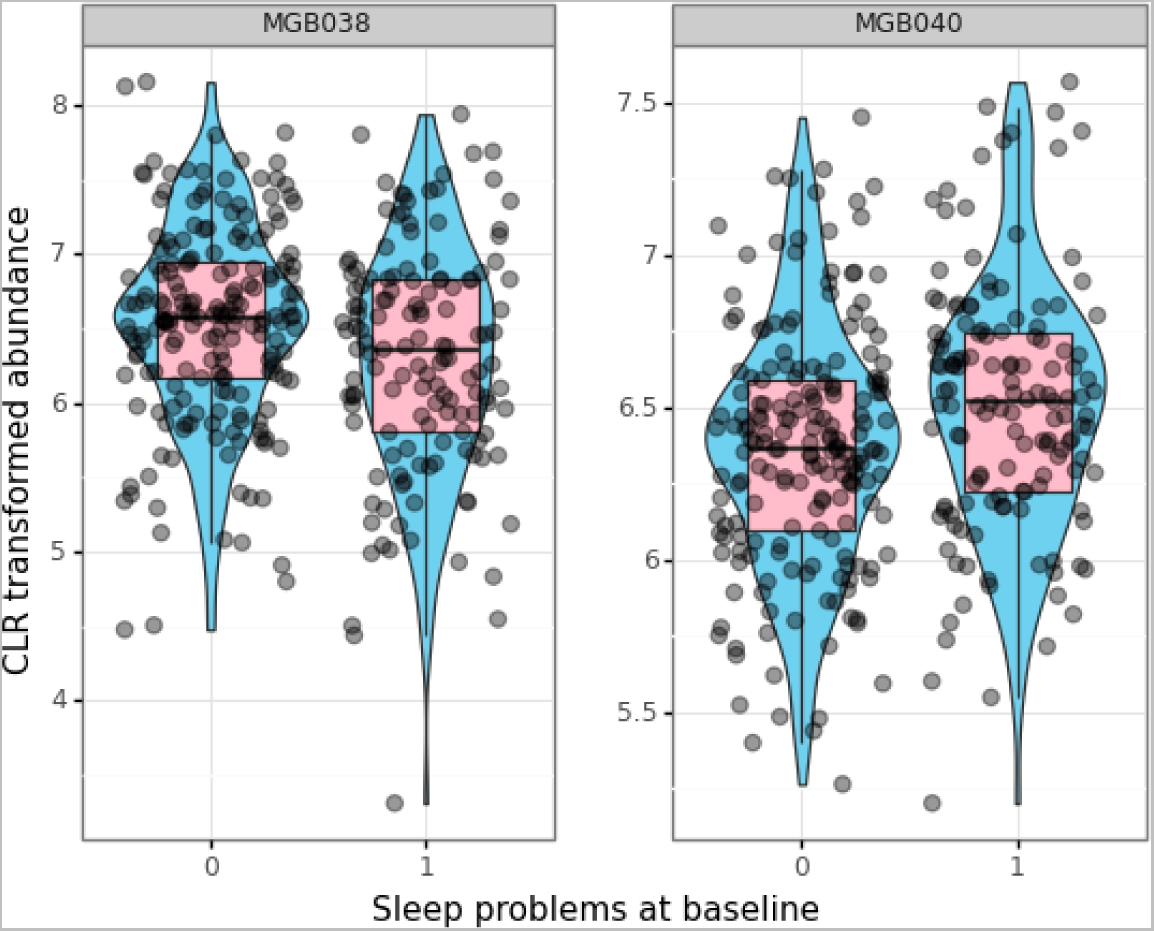
Boxplot of microbiome features associated with sleep problems at baseline. Each panel provides violin plots (sky blue fill), box plots (pink fill), and horizontally jittered points presenting CLR transformed abundances of bacterial genera and functional pathways (y-axis) for each level of the outcome measuring the reported sleep problems at baseline: yes = 1 and no = 0 (x-axis). Panel titles display the name of the respective microbiome feature.

**Figure S10.**
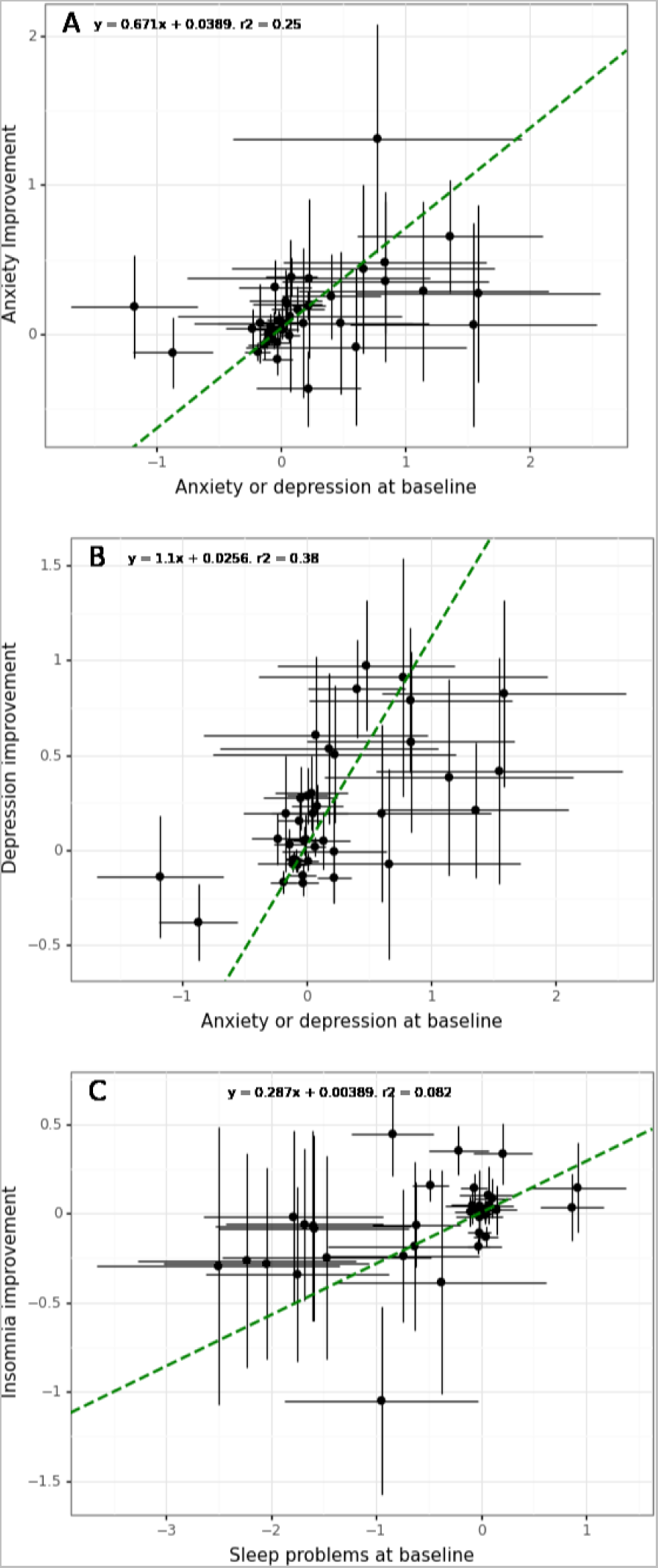
Comparison of functional pathway associations with outcomes at baseline versus improvement at follow-up. X and Y-axis provide regression coefficients from the models fitted for anxiety or depression, or sleep problems at baseline versus improvement in anxiety (panel A), depression (panel B) or insomnia (panel C) at follow-up with the corresponding standard errors. Dashed line is the line fitted using orthogonal distance regression that accounts for the standard error on x and y variables. If there was a perfect correlation between the variables, the points would lie on the diagonal and the r-squared would be 1. We used a linear model of type y = x*b + a and each panel presents regression coefficients and r-squared values.

**Figure S11.**
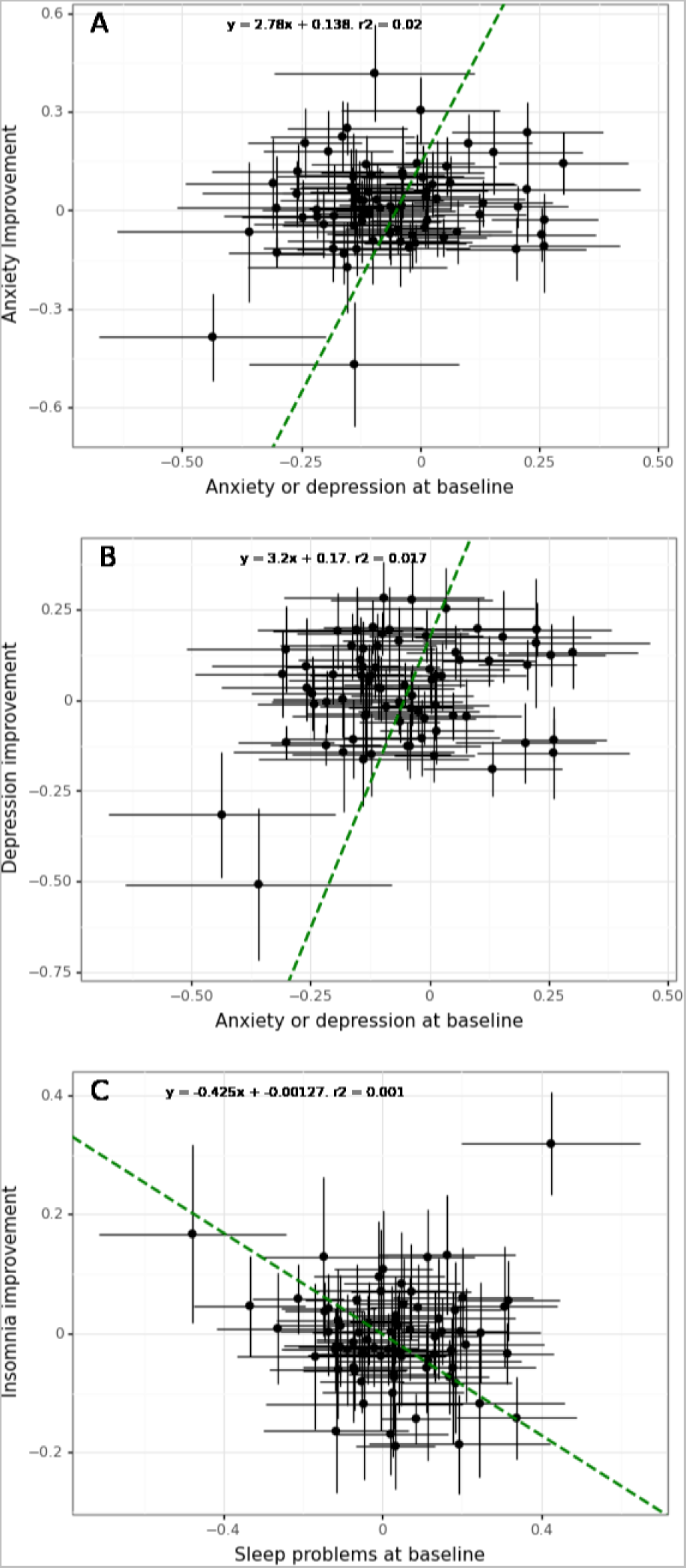
Comparison of bacterial genera associations with outcomes at baseline versus improvement at follow-up. X and Y-axis provide regression coefficients from the models fitted for anxiety or depression, or sleep problems at baseline versus improvement in anxiety (panel A), depression (panel B) or insomnia (panel C) at follow-up with the corresponding standard errors. Dashed line is the line fitted using orthogonal distance regression that accounts for the standard error on x and y variables. If there was a perfect correlation between the variables, the points would lie on the diagonal and the r-squared would be 1. We used a linear model of type y = x*b + a and each panel presents regression coefficients and r-squared values.

**Figure S12.**
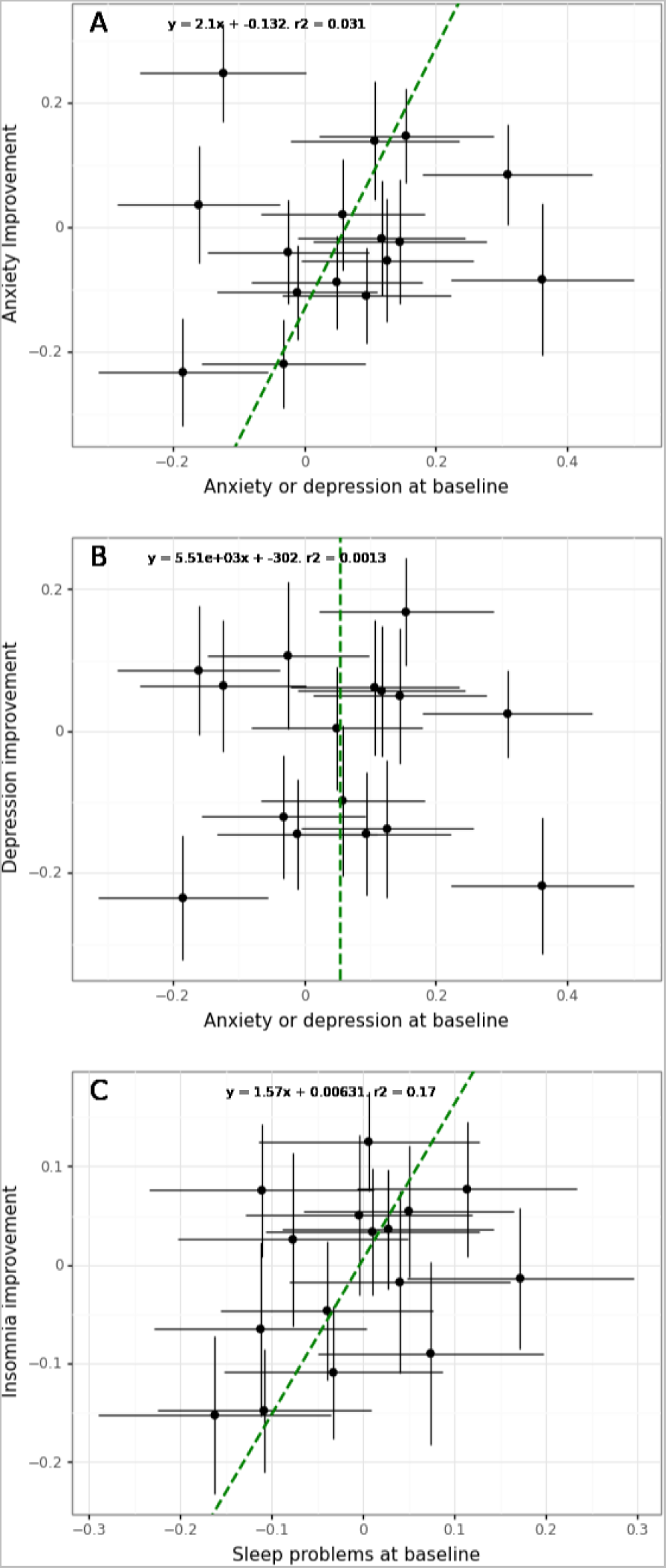
Comparison of genetic scores associations with outcomes at baseline versus improvement at follow-up. X and Y-axis provide regression coefficients from the models fitted for anxiety or depression, or sleep problems at baseline versus improvement in anxiety (panel A), depression (panel B) or insomnia (panel C) at follow-up with the corresponding standard errors. Dashed line is the line fitted using orthogonal distance regression that accounts for the standard error on x and y variables. If there was a perfect correlation between the variables, the points would lie on the diagonal and the r-squared would be 1. We used a linear model of type y = x*b + a and each panel presents regression coefficients and r-squared values.

